# The effectiveness of surveillance technology for the prevention of suicides in public spaces: a systematic review

**DOI:** 10.1101/2025.10.17.25338217

**Authors:** Laura Joyner, Bethany Cliffe, Jay-Marie Mackenzie, Keith Hawton, Peter Craig, Lisa Marzano

**Affiliations:** Department of Psychology, Middlesex University, London, UK; Department of Psychology, University of Westminster, London, UK; Department of Psychiatry, Centre for Suicide Research, University of Oxford, Oxford, UK; School of Health & Wellbeing, University of Glasgow, Glasgow, UK

**Keywords:** Suicide, Suicide Prevention, Surveillance Technology, CCTV, Systematic Review

## Abstract

**Background:** The use of surveillance technologies has been recommended for supporting suicide prevention efforts in public spaces. We sought to identify and synthesise the current evidence on the impact of surveillance technologies deployed in such environments on suicide and related outcomes.

**Methods:** We conducted systematic searches between 1990 up to August 2025 on the following databases: PsycINFO, MEDLINE, CINAHL, Computer Source, Ovid, SPP, Web of Science, PTSDpubs, CENTRAL, ACM DL, and IEEE Explore. Our searches also extended to grey literature from relevant websites. Studies were included if they assessed the deployment of surveillance technology at a public location on suicide-related outcomes (including suicides, suicide attempts, rescue interventions, and trespass events). Study results were synthesised using narrative and tables.

**Results:** The searches identified 2,647 items, with 15 studies meeting the eligibility criteria. Just one study provided clear evidence of a reduction in suicides at a site (a bridge) after installation of a “smart” surveillance system, but its use of tension-wire sensors also restricted ease of physical access. The presence of CCTV cameras was associated with lower rates of suicide at local rail stations in one study, but this was not a significant factor in two other studies of metro stations. Two studies observed an increase in interventions following the installation of smart surveillance technologies (SSTs) on bridges, but with no change in suicides rates. Three studies indicated that SSTs with audible deterrents may reduce trespass to dangerous sites, but their relevance to suicide prevention is unclear.

**Conclusions:** The current evidence base to support the use of surveillance technologies for preventing suicides is limited. There is a clear need for further evaluations of surveillance-based interventions, including their implementation, associated measures and impacts to inform development of effective suicide prevention initiatives.

**Key messages:** *What is already known on this topic:*

- It is thought that surveillance technologies may aid suicide prevention efforts at public locations by increasing opportunity for human intervention. However, it is unclear whether they are effective at preventing suicides.

*What this study adds:*

- ur systematic review found 15 studies that examined the effectiveness of installing surveillance technologies at preventing suicides or trespass at public locations.
- verall, the evidence of surveillance technologies helping to prevent suicide was limited.
- some locations the installation of surveillance technologies was associated with an increase in numbers of interventions or police call outs, but not a decrease in deaths.
- use of audible deterrents may be effective for preventing trespass, however, to date there remains no evidence that they will help to prevent suicides.

*How this study might affect research, practice or policy:*

- The findings highlight critical gaps in the current evidence, particularly in regard to the use of “smart” or “intelligent” surveillance technologies.
- Further research is also needed to understand the impact of processes surrounding surveillance technologies on their effectiveness (e.g., rescue response times).

## INTRODUCTION

Approximately 7,000 suicides are recorded annually in the United Kingdom (1), with around a third of these deaths occurring in public locations, such as railways, cliffs, roads and bridges (2). These deaths can have a lasting impact, not only on family and friends, but also for potential witnesses and staff dealing with their aftermath. There can also be substantial financial implications for affected sectors. For example, each suicide on the railways in Great Britain is estimated to cost the industry approximately £275,000 (3).

Strategies to prevent suicides in public locations are often categorised into four general approaches: (1) restricting access to the site and means of suicide, (2) increasing opportunity and capacity for human intervention, (3) enabling help-seeking, and (4) changing the public image of the site (4). While measures that fully-restrict access to means (e.g., installing barriers over 2.3 metres) have proven effective in preventing suicides in some environments (5), such approaches are not always practical or feasible to install everywhere (e.g., due to engineering constraints). Another “hard” measure recommended in national guidance is the introduction of surveillance technologies (4).

Previously, authors of systematic reviews of suicide prevention measures at high-risk locations have discussed surveillance cameras in the context of “increasing opportunity for human intervention” (6–8). However, others have noted that such systems require on-going monitoring by vigilant staff to identify potential suicide attempts in real-time (9) and therefore their use may not always be practical. However, by integrating alternative data sources (e.g., heat sensors) and / or tools (e.g., analytics to detect motion in a scene) “Smart” Surveillance Technologies (SST) can automate aspects of the surveillance process in ways that may alleviate some (or all) of the monitoring burden for staff (10). In some cases, they may also be designed to issue a fully-automated response (e.g., audible deterrents, responsive lighting). SSTs have been proposed, recommended, and utilised in suicide prevention efforts for some time (11,12), and are increasingly becoming more complex, technologically advanced, and commercially available. Examination of if, and when surveillance technologies (including SSTs) may help prevent suicides in public locations is warranted in order to inform future suicide prevention initiatives. The aim of this paper is therefore to examine the existing literature on the effectiveness of implementing surveillance technology at preventing suicides in public spaces.

## METHOD

The review protocol was registered (PROSPERO ID CRD42024495308) on 17^th^ January 2024. This work was part of a wider review looking at the use of surveillance in suicide prevention at public locations. However, for clarity, other objectives from this review (e.g., feasibility) will be explored elsewhere.

### Eligibility Criteria

For the present work key outcomes included: 1) deaths by suicide, 2) suicide attempts, 3) intervention events (callouts, human intervention, calls to telephone hotlines, etc.). In line with previous work (e.g., Havârneanu et al., 2015), outcomes related to trespass were also included as an associated and preventable high-risk event (for the objective of the present review only). Only examples of surveillance technologies being used to monitor behaviour within public spaces (e.g., railways, bridges, coastal locations) were included.

Synthesis was initially grouped into themes according to the research questions and type of automated detection employed by the system. These were: “No smart monitoring” (e.g., CCTV), “detect presence” (e.g., motion sensors), “detect specific action” (e.g., linger detection), “detect emotional states”, “Identify specific people”, and “forecasting risk of attempt”. Coding of these modalities was based on the behaviours and / or characteristics the SST sought to detect and may therefore differ from formal technological or author descriptions. Where a single study employed more than one modality, categorisation was prioritised based on whichever was the more advanced category. For instance, an intervention employing both intrusion detection and behavioural detection would have been categorised under the latter.

### Identification of eligible studies

The following databases were searched: PsycINFO, MEDLINE, CINAHL, Computer Science, Ovid, SPP, Web of Science, PTSDpubs, CENTRAL, ACM DL, and IEEE Explore. Searches also extended to grey literature, including government, business, industry and third-sector reports and non-peer reviewed academic work. The full search strategy can be found in Appendix A. An initial search was run in March 2024, with follow-up searches in October 2024 and August 2025 to ensure publications in this base review were up to date. Any updates from subsequent searches during the course of the project will be published in a public OSF project (https://osf.io/jvmxf/overview?view_only=d0636100198b44cc9328495025ce94a9). No restrictions were applied regarding publication type. Only studies published in English since 1990 were included.

After the initial round of searches was completed, files containing search results were uploaded onto Covidence for deduplication. Two researchers (LJ and BC) independently screened titles and abstracts on Covidence and crosschecked for agreement. The same reviewers then independently screened the remaining full-text articles, documenting reasons for exclusion at this stage. Disagreements were discussed and resolved between the two reviewers.

### Data Collection & Quality Assessments

A data extraction tool developed using Microsoft Excel was piloted prior to data capture. Two researchers (LJ & BC) extracted data from papers and cross-checked 10% of extracted data to ensure accuracy. Any disagreements unresolvable by discussion were referred to a third reviewer (JM or LM). An overview of extracted data can be found in Appendix B. Quality appraisals were carried out using the Mixed Methods Appraisal Tool (MMAT) (14). Studies were first checked for eligibility using the MMAT’s two screening questions, and eligible studies were rated against five criteria from the Quantitative non-randomized category. Ratings of “Yes” were assigned where studies reported information demonstrating they had met the criterion, otherwise they were given ratings of “No” or “Can’t Tell”. The quality appraisals were conducted by one reviewer (LJ) and checked for accuracy consistency by a second (BC), with discrepancies resolved by discussion (Appendix C).

### Data Synthesis

Extracted data were synthesised using narrative and tables. Meta-analysis was not conducted as data from the relevant studies were not sufficiently homogenous.

### Involvement of Lived Experience Experts

People with lived experience of suicide have been involved at several key stages of our project. Lived Experience Experts from the National Suicide Prevention Alliance (NSPA) were consulted prior to making the funding application associated with this work, and provided feedback on the project’s aims and objectives, methods, and dissemination plans. During the course of the project, we have continued to meet regularly with our Lived Experience Advisory Group (LEAG), whose feedback has helped us to make sense of some of the potential risks surrounding the different technological responses, for example. A member of the LEAG also sits on our project steering group where, as a direct result of their feedback, lived experience feedback and consultation are included as a prominent agenda item.

## RESULTS

After removal of duplicates, a total of 2,647 items were identified by 16^th^ October, 2024. After initial screening of titles and abstracts, 296 items were assessed for eligibility. Of those, 118 were excluded at this stage, with reasons for exclusion provided in Figure 1. A total of 178 documents included discussion of the role of surveillance technologies in suicide prevention. However, only 15 of these fulfilled the criteria for inclusion in the present work (i.e., looked at effectiveness). No additional studies for this review were identified from follow-up searches between 21^st^-27^th^ August 2025.

**Figure 1.**
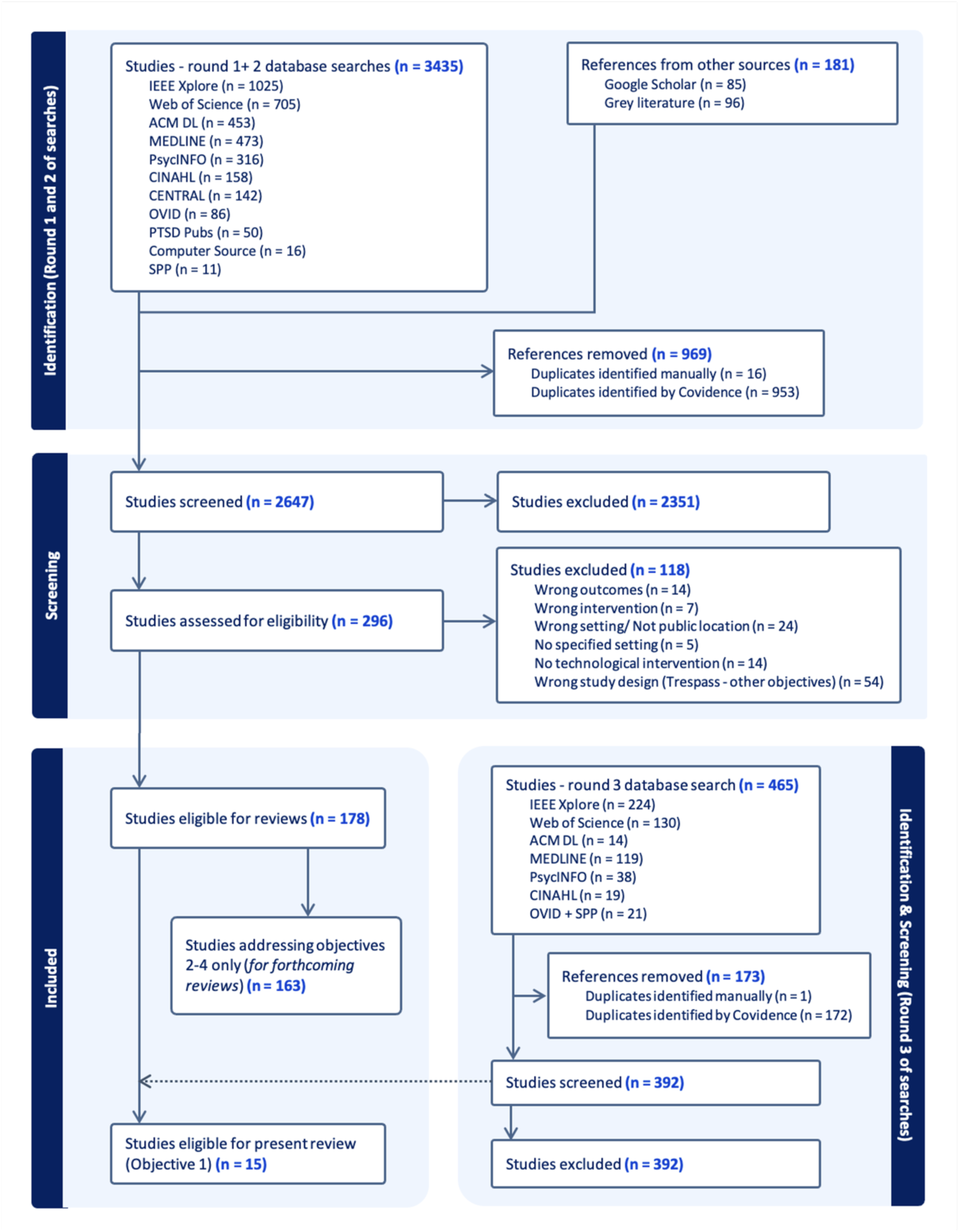
Preferred Reporting Items for Systematic Reviews and Meta-Analysis (PRISMA) flowchart

### Study characteristics and quality appraisal

The included studies came from eight countries: Australia (*n* = 5), South Korea (*n* = 4), Austria (*n* = 1), Belgium (*n* = 1), Denmark (*n* = 1), Finland (*n* = 1), Sweden (*n* = 1), and the United States of America (USA: *n* = 1). Three of these studies examined the same project implemented at a coastal location in Australia, each at a different point in time. A further five studies focussed on bridge locations, with potentially up to three having investigated different interventions deployed on the same bridge in South Korea. Another seven studies focussed on railway or metro locations, and, of these, four were of suicide-related outcomes at stations and/or their surrounding environments. The remaining three rail (trespass-focused) studies examined systems installed at a rail bridge, a tunnel entrance, or at lineside locations.

Eleven of the 15 studies included deaths by suicide as an outcome, with four also analysing suicide attempts; however, this outcome was not clearly defined across all studies. Single studies investigated intervened suicidal acts, police call outs and successful sightings on CCTV, and at number of rescue attempts. Three studies had trespass events as the main outcome.

The quality appraisals of the included studies are reported in Appendix C. Studies achieved between 0%-60% of the MMAT quality criteria for quantitative non-randomized studies. It is important to note, however, that study ratings were affected due to MMAT Q.3.1 which asks about representative of participants, which in the case of these studies was not appropriate. Moreover, five of the studies identified changes to the way in which the systems operated (e.g., periods where the SST was not operational, upgrades to the system), and it was unclear whether such changes had occurred during the data collection periods for the other studies. Overlap with the installation of other mitigating initiatives (either as part of the same study or reported elsewhere) meant that confounders could not be accounted for in several of the studies. Also, two studies did not provide sufficient information to determine the appropriateness of measurements or completeness of outcome data.

### Characteristics of the interventions

Summaries of individual studies can be found in Table 1. Nearly half of the studies included the use of systems that we primarily categorised as “not smart” e.g., CCTV (7/15). These included one study where CCTV was monitored by security officers at the locations as part of their role (rather than the system playing a role in identification or triggering a response). Another CCTV system installed at one location and examined in three studies was initially monitored upon police request only (by a security company), however, “smart” upgrades during the post-intervention period for two of these studies meant that the system later issued automated alerts when barriers were climbed. However, in three other studies it was unclear whether the CCTV systems were, or could, be monitored in real-time.

**Table 1.**
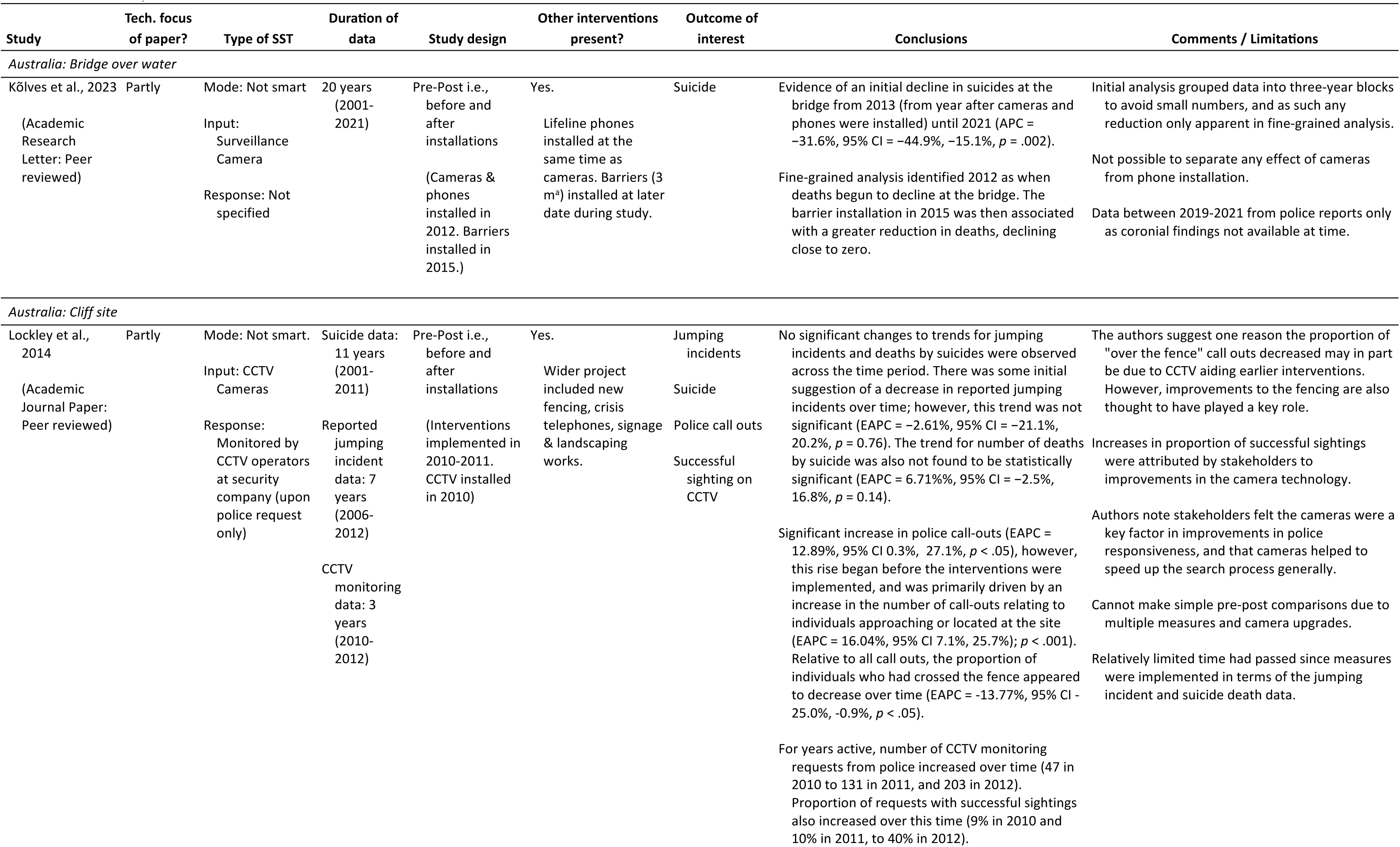
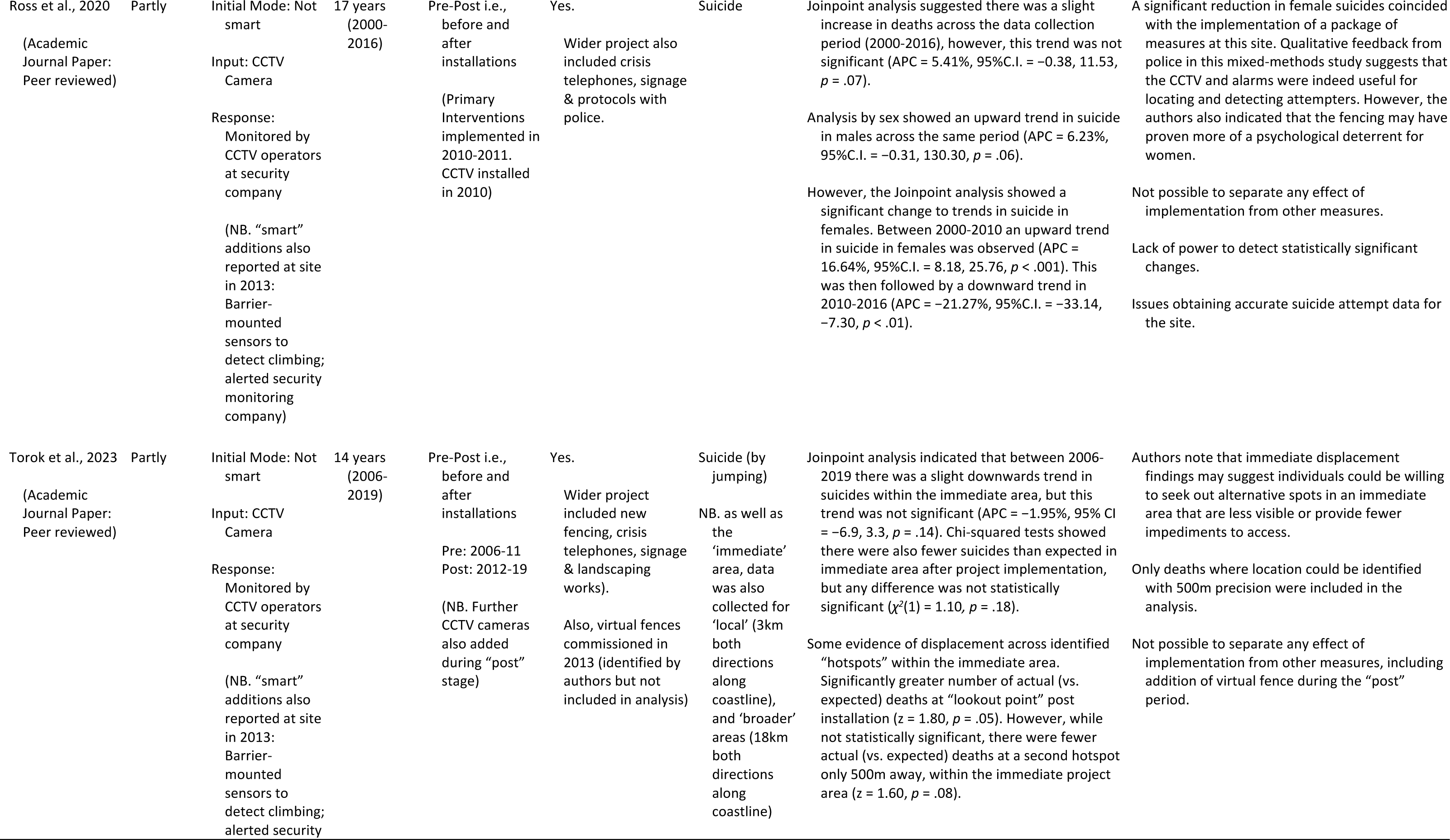
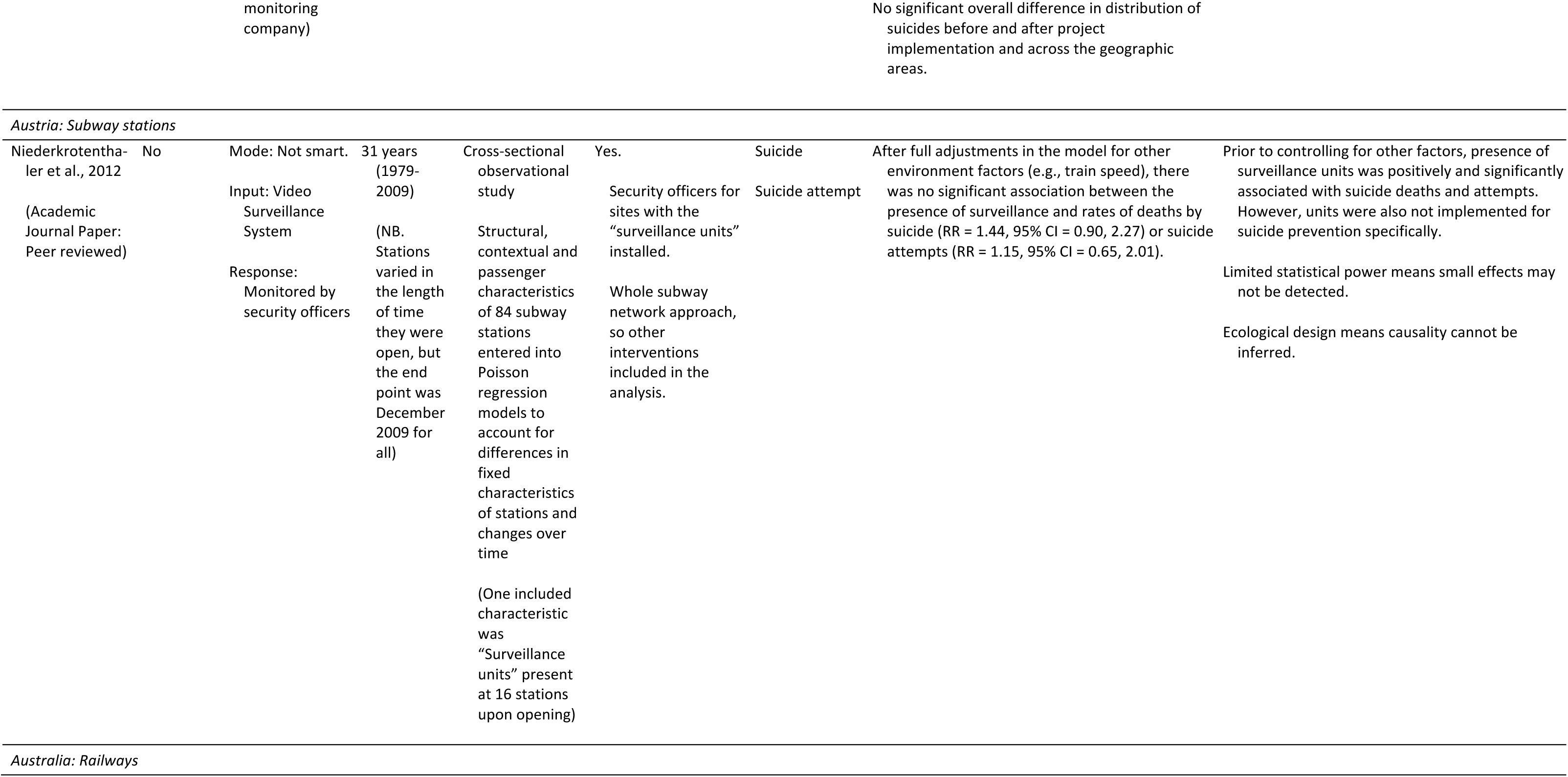
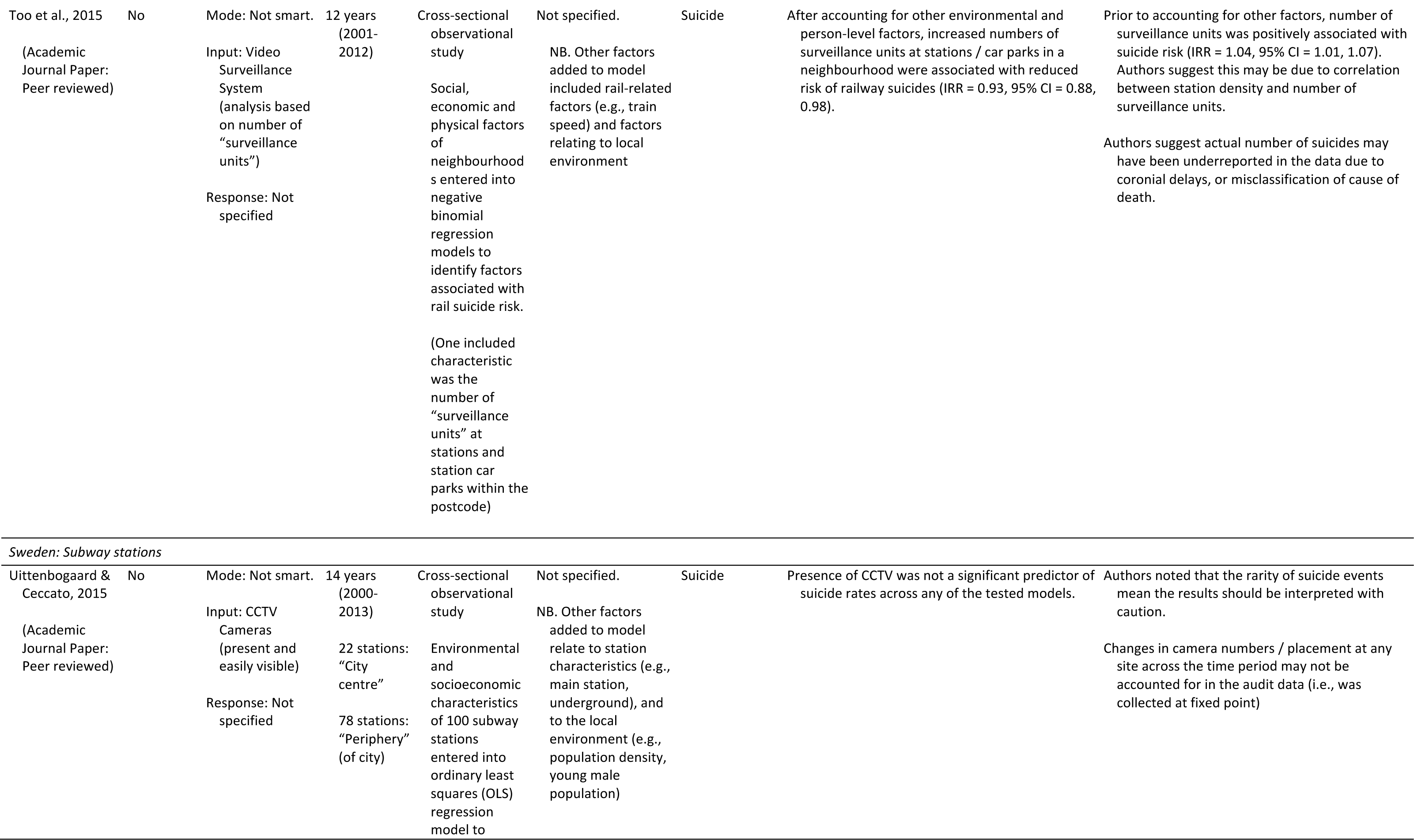
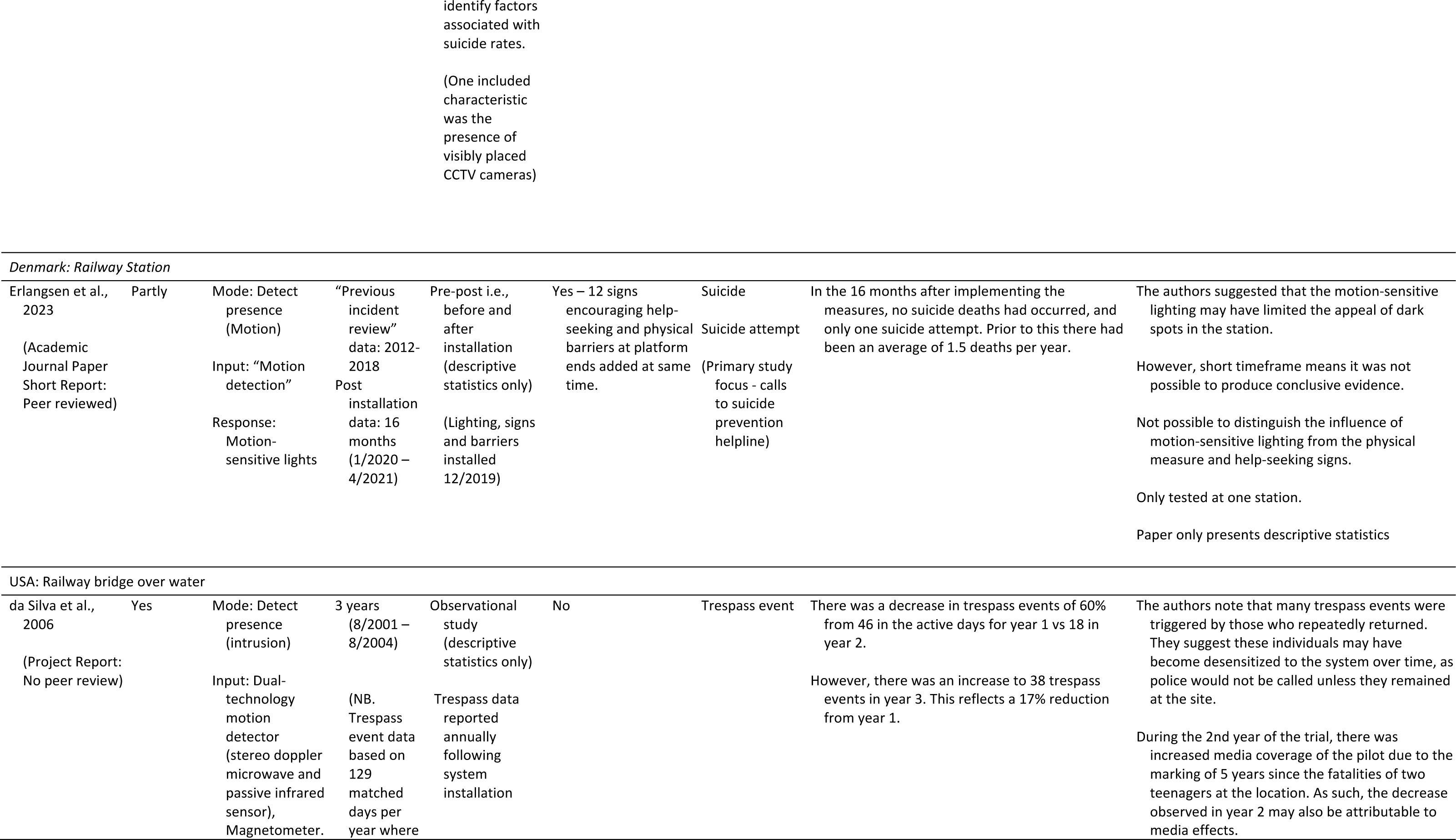
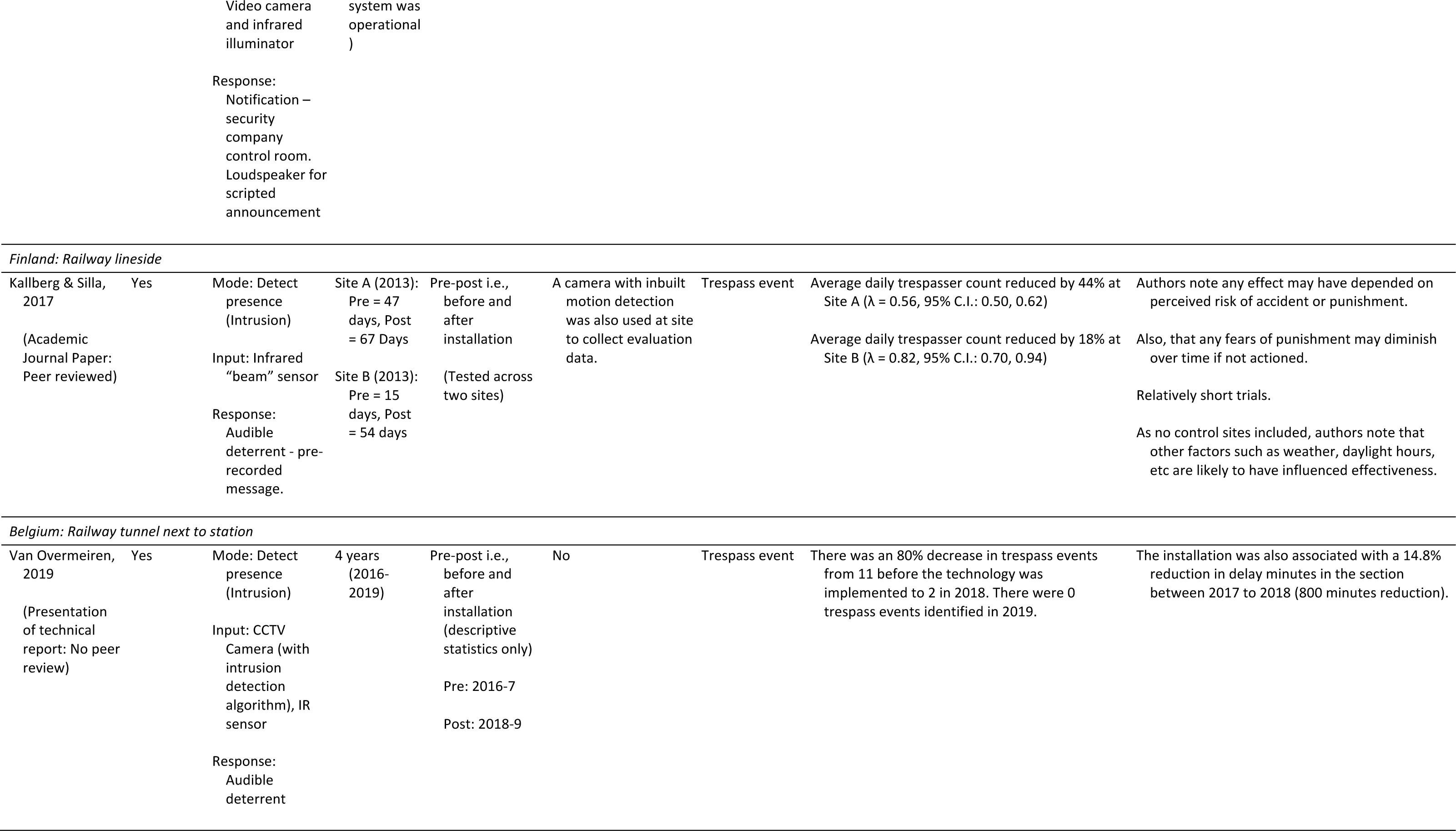
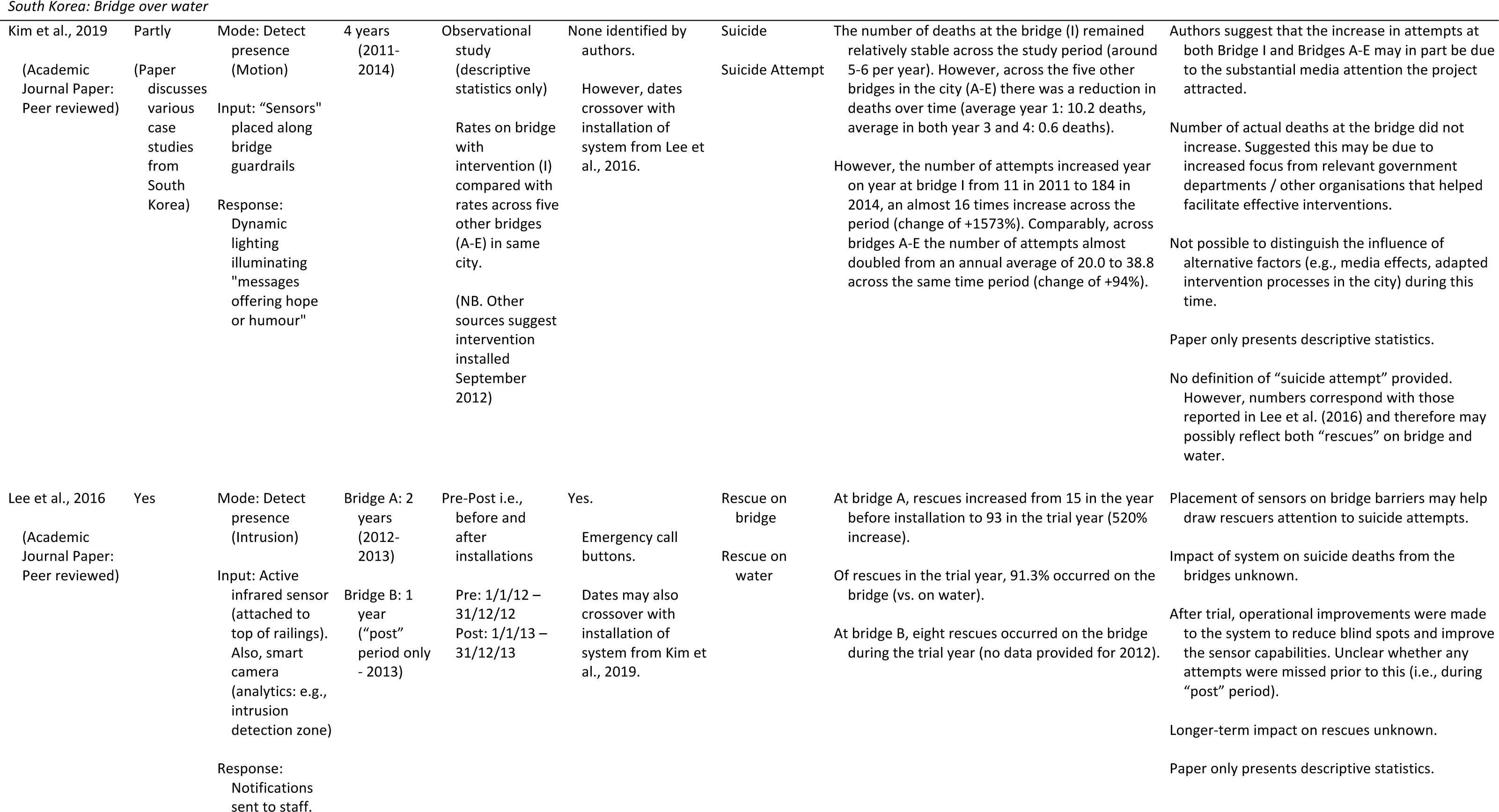
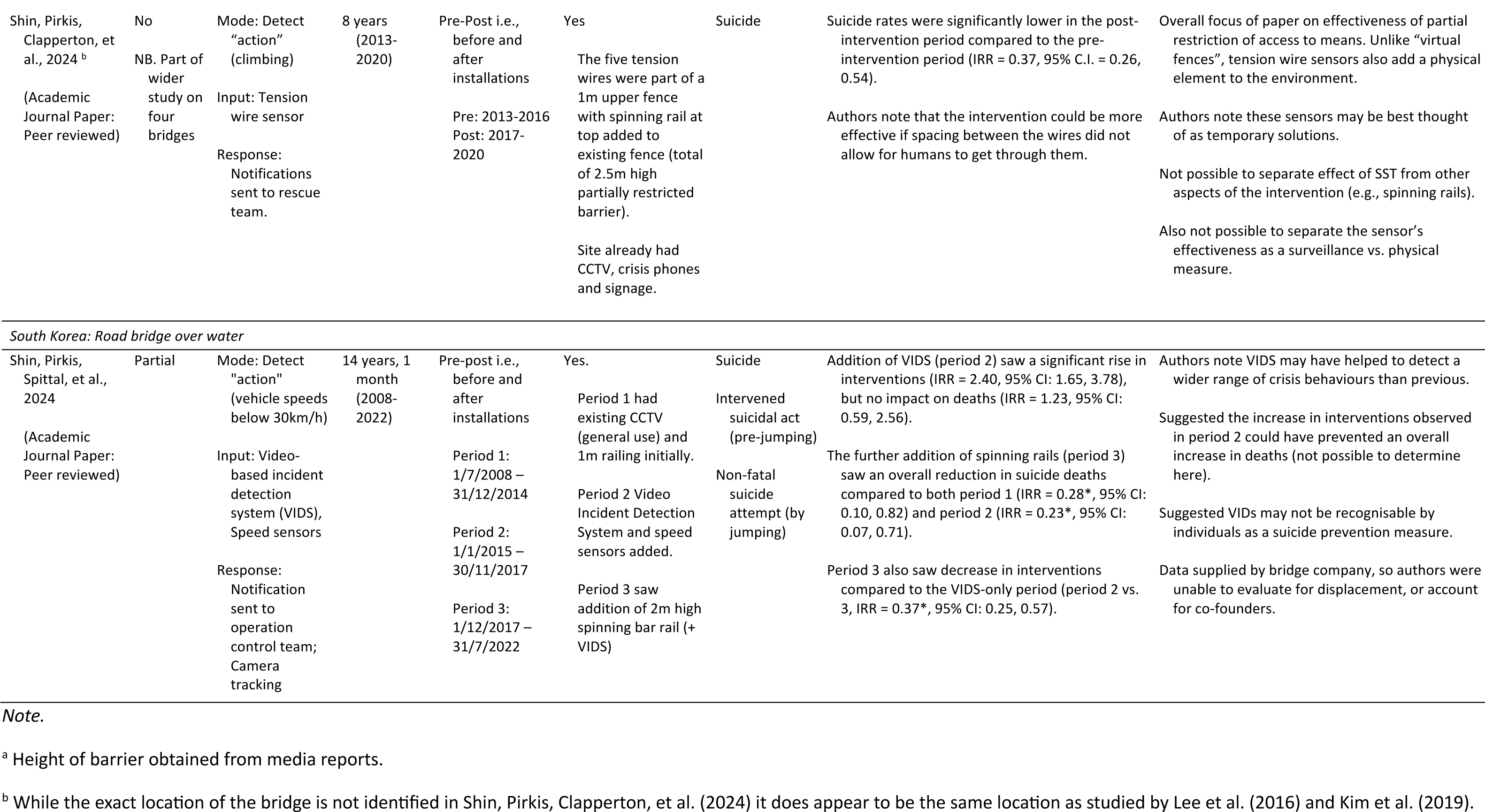
Summary of studies.

A further six studies focussed on interventions which included the use of “smart” surveillance technologies (SSTs) designed to “detect the presence” of an individual within or entering a region of interest. The remaining two studies examined interventions which included SSTs that we classed as detecting a specific “action” (e.g., vehicle slowing, climbing). To date, we have identified no studies which have investigated the effectiveness of any SSTs designed to “detect emotional states”, “identify specific individuals”, or “forecast risk of attempt” (e.g., based on exhibited behaviours) at reducing suicide or trespass-related outcomes.

Across the studies, four included SSTs designed to operate as stand-alone interventions (i.e., fully automated), two of which were tested in relation to trespass outcomes only. Another study tested an SST which notified staff who were then to use the system’s speakers to issue a scripted trespass intervention (i.e., technology mediated intervention). The remaining three studies focused primarily on SSTs designed to notify staff who would then initiate a rescue response (i.e., human-led intervention).

#### No “Smart” Monitoring

We identified seven studies where surveillance cameras were included in natural experiments (Table 1). Of these, three included video surveillance units as one of several “station characteristics” as potential predictors of suicide rates across railway or subway networks. Niederkrotenthaler et al. (18) examined the influence of different station characteristics on suicide attempts and deaths between 1979-2009. This included the presence of surveillance units at 19% of stations on the Vienna subway network (16/84 stations), consisting of a video surveillance system monitored by officers (who also conducted patrols) installed for general use when the station opened. Prior to adjustment, Poisson regression models suggest the presence of surveillance units was one factor positively associated with rates of suicide deaths and suicide attempts. However, after adjusting for covariates such as speed of train and passenger numbers the presence of surveillance units was no longer significantly associated with rates of suicide deaths or suicide attempts.

Similarly, Uittenbogaard & Ceccato (19) included several station environment characteristics in their analysis of suicide deaths between 2000-2013 on the subway system in Stockholm, including the visible presence of CCTV. Ordinary least squares regression models showed no significant association with the presence of CCTV on suicide rates at stations across the total subway system, or at 78 periphery stations. Due to multicollinearity, presence of CCTV was not included in the city centre model (22 stations). It is also unclear whether the CCTV was being monitored in real-time.

Too et al. (20) looked at the influence of various social, economic, and physical factors on rail suicide rates across neighbourhoods in Victoria between 2001 and 2012. This included the number of video surveillance units installed at railway stations and car parks within the local area. Univariate analysis suggested a positive relationship between suicide risk and the number of surveillance units within a local area. However, in the multivariate model the relationship became negative, which the authors suggest may be due to correlations between station density and the number of surveillance units. While it is unclear whether the system was monitored in real-time, this approach may better reflect the overall (perceived) coverage of surveillance than the previous studies in station environments.

A recent study by Kõlves et al. (21) analysed suicide data between 2001 and 2021 from one Australian bridge location to understand the effect of different packages of suicide prevention measures installed across time. Prevention measures included the installation of crisis phones, signage, and surveillance cameras in 2012, and barriers in December 2015. To avoid small numbers the authors also grouped data into 3-year blocks. Using Joinpoint regression analysis, they identified two changes in trends across the data collection period, including a decrease in deaths by suicide at the bridge from 2013 onwards. While descriptive statistics would indicate no change between the three years prior to (2010–12) and after the installation of cameras and phones (2013–15), the authors suggest 2012 was the start of the decline (but deemed the subsequently installed barrier to be more effective at preventing suicides). The authors also found no evidence of substitution of deaths to other locations. Furthermore, during the same time period in the surrounding areas observed trends in suicide numbers either increased or remained somewhat stable, suggesting the reduction in deaths at the bridge was not due to wider trends in suicide at the time. A further three studies in Australia examined the influence of implementing a package of measures (including CCTV, and also fencing, crisis telephones, signage and landscaping works) as part of a large suicide prevention project at one coastal location (22–24). The latter two studies also mention a sensor-based “virtual fence” installed approximately three years after the core package of measures, including the CCTV, which were the focus of these studies (however, the installation of the virtual fences does fall within these study timeframes). None of the studies reported a significant reduction in suicide deaths in the periods after CCTV installation (i.e., two-three years (22), six years (23), or up to nine years (24)). However, in the most recent study exploratory kernel density analysis indicated that, after the project had been implemented, the distribution of the number of suicides occurring at specific areas within the immediate environment may have changed (24). Specifically, one area within the environment that had been associated with the greatest number of deaths before the measures were implemented (actual *n* = 41, expected *n* = 36) had fewer than expected deaths in the period after (actual *n* = 34, expected *n* = 38), although this change was not significant. There was an increase in deaths at a second location in the same park after the measures were implemented (pre *n* = 9, post *n* = 18), and this was greater than expected (expected values: pre and post *n* = 13). The authors noted that the close proximity of both sites may suggest immediate displacement, and that some people may have been willing to find areas with reduced visibility. However, due to data sparseness they were unable to conduct inferential spatial analysis. Wider trends across the city and broader area were also taken into account by the authors (24).

Lockley et al.’s (22) study also identified a significant increase in the overall number of police call-outs at the same location over time, but noted that the proportion of episodes where an individual was located “over the fence” was significantly reduced. They suggested that may be likely due to fence improvements but acknowledged that the cameras may have also played a role in aiding earlier intervention. Indeed, as the CCTV system was improved over time, both the number and proportion of successful CCTV assistance requests from police increased (from 9% in 2010 to 40%). This may suggest that the CCTV system supported rescue responses in some cases.

#### Detect “presence” (e.g., motion, intrusion, obstacle)

Across the literature, a range of technologies have been used or proposed to aid the detection of individuals within an environment or crossing a virtual boundary. Such systems may integrate sensors or apply analytics to detect changes within an environment (e.g., introduction of body heat, new objects within a region). Unlike non-smart surveillance technologies (e.g., CCTV cameras) which rely on continuous real-time monitoring, smart surveillance technologies may produce automated, technological responses at the site itself (e.g., audible deterrents) instead of, or in addition to, notifying officials (e.g., control room staff).

We identified six studies testing the effectiveness of systems designed to detect the “presence” of a person: three focusing on suicide-related outcomes, and three for trespass more broadly. Four of these studies focused on railway environments, specifically; stations, rail-bridges, tunnels, and lineside. Two further studies investigated different SSTs installed on bridges with pedestrian access; however, both included data from the same bridge and time period (16,17).

##### Notify for human-led response

One study in South Korea evaluated a system designed primarily to alert control centre staff and was piloted on two river bridges throughout 2013, comprising primarily an infrared sensor and smart camera with intrusion detection zone (16). One of the bridges was reported to have seen a 520% increase in rescues: from 15 in 2012 to 93 in 2013 (including eight on the water). However, due to the cross-over with installation dates of another pilot on this bridge (i.e., Kim et al. (17)) it is not possible to attribute any change in rescue numbers to this project.

##### Activate lighting

We also identified two studies which included SSTs which activated lighting-based interventions. In one study investigating the installation of a package of measures, including motion-sensitive lighting, signage and platform end barriers, at one railway station, Erlangsen et al. (25) reported that no deaths had occurred in the 14 months after installation. The years prior to installation (2012–18) had seen an average of 1.5 deaths per year, sometimes at secluded parts of the platform. The authors were unable to produce conclusive evidence regarding effectiveness due to the short timeframe, nor would it be possible to attribute effects to any single measure.

In another study examining a variety of different suicide prevention measures, Kim et al. (17) present the 2011-2014 descriptive data around the “Bridge of Life” project (deployed at the same bridge in South Korea as the pilot reported by Lee et al. (16)). This project, installed in the latter half of 2012, used motion detection sensors to dynamically illuminate “messages of hope”. As previously noted, suicide attempts at the bridge increased from 15 in 2012 to 93 in 2013, and then 184 in 2014. However, in marked contrast, the number of deaths by suicide at the site appeared to remain stable (6 in 2012, 5 in 2013, 5 in 2014). The authors suggest that the increase in attempts may be in part due to the substantial media attention towards the project. Indeed, descriptive statistics summarising events from across five other local bridges suggest attempts remained relatively stable between 2011 and 2013 before almost doubling in 2014.

##### Audible deterrents

No studies identified to date have examined the effectiveness of using announcements or “audible deterrents” on suicide-related outcomes. However, we identified three reports of initiatives where such a system was tested against trespass-related outcomes, all on the railways. Firstly, da Silva et al. (26) reported findings from a system trialled in the USA between 2001-2004 that notified a security control room when trespassers were detected on a railway bridge. Staff then activated the loudspeaker to make a scripted announcement requesting trespassers leave the area. Da Silva et al. (26) reported a 60% reduction in trespass from year 1 (46 events) to year 2 (18 events), and then a subsequent somewhat smaller increase in year 3 (38 events). The authors suggest that this rise may have been due to desensitisation over time in some individuals who trespassed repeatedly without consequence (e.g., police intervention). However, they also noted that there had been media coverage in year two about the project and prior fatalities at the location.

Another project tested the effectiveness of an automated audible deterrent installed at two lineside locations on the railways in Finland for approximately two months (27). At Site A, the installation of the system was associated with a 44% reduction in daily trespass (λ = 0.56, 95% C.I.: 0.50, 0.62). Site B saw an 18% reduction (λ = 0.82, 95% C.I.: 0.70, 0.94). The authors noted that the difference may have occurred because Site A had a higher daily number of trains than Site B (120 vs. 21), which also travelled at a faster speed (up to 120km/h vs 50km/h). As such, the perceived risk potential of trespassing could have been greater at Site A than for Site B, which may explain the differences in effect sizes.

Finally, Van Overmeiren (28) described a piloted system which issued audible deterrents when trespassers were detected at a railway tunnel in Belgium. Descriptive statistics suggest there had been 11 trespass events in the two years prior to installation. The year after installation saw two trespass events, and the subsequent year saw none. The installation was also associated with a reduction of 800 delay minutes (−14.8%) for trains in the area from the year prior to installation compared to the first year.

#### Detect specific “actions”

We identified two studies which tested the effectiveness of suicide prevention measures which included SSTs designed to detect specific “actions”, both being at bridge locations and intended to alert a rescue team.

Using data from a road-bridge in South Korea (2008–2022), Shin, Pirkis, Spittal, et al. (29) investigated the effect of installing a Video-based Incident Detection System (VIDs) in 2015 which utilises CCTV cameras and speed sensors to detect vehicles travelling below 30km/h, and also the addition of a 2m high spinning bar rail in late 2017 (previous to these upgrades, the bridge had only a standard 1m rail and CCTV system). Incident rate ratios (IRR) indicated a significant increase in interventions following the installation of VIDS (IRR = 2.40, 95% CI: 1.65, 3.47), but this reduced following the installation of the barrier rails (IRR = 0.37, 95% CI: 0.25, 0.57). There was no significant effect of installing VIDS on the number of deaths by suicide (IRR = 1.23, 95% CI: 0.59, 2.56), but there was again a reduction in deaths after barrier rails were installed (IRR = 0.23, 95% CI: 0.07, 0.71). The author suggest that VIDS may allow atypical behaviours to be identified that could have otherwise been missed, potentially including individuals experiencing different degrees of crisis.

Another study examined a variety of partial restriction measures across four bridges in three countries, including a site where in late 2016, a 1 metre upper barrier of five tension wire sensors topped with a spinning rail had been installed atop of an existing 1.5 metre barrier (15). While the five wires had spaces between them, attempts to climb (e.g., by pulling a wire by 10cm or more) or cut the wire sensors would lead the SST to alert a rescue team. Using data from 2013-2020, the authors reported a significant decrease in the number of suicides in the period after installation (IRR = 0.37, 95% CI: 0.25, 0.54). They suggested that this may be because the wires and spinning rail make access more difficult. As intervention-related outcomes were not reported, it is difficult to ascertain the extent to which the sensors may have also played a role in the positive outcome (e.g., automating notifications of climbing attempts to rescuers).

## DISCUSSION

In the searches for this review, we identified 15 studies which looked at whether the presence or installation of surveillance technologies (often alongside other measures) influenced rates of suicide or related events at public locations. Overall, the evidence provided by these studies is mixed. Increases in interventions or rescues were identified in two studies after SSTs were installed on bridges to detect vehicles slowing down (29) and people leaning over railings (16). Elsewhere, a package of measures installed at a coastal location (including a 1.3 m barrier and a CCTV system) was also associated with an overall increase in police call outs, a decreasing proportion of which involved “over the fence” call outs (22). Given that surveillance technologies are often categorised as a measure to help increase opportunity for human intervention, these associations may be somewhat expected. Despite the associated increase in rescues / interventions in these studies, however, only one reported a significant reduction in deaths. This was not associated with the installation of the SST, however, but the subsequent addition of a 2-metre barrier (29). Similarly, while Kõlves et al. (21) suggest that the decline in suicides at one bridge site in Australia began following the installation of cameras and crisis phones, there was a much clearer reduction in deaths following the installation of 3-metre barriers at the site three years later.

Across the review there was only one study identified where the introduction of measures, including an SST, was statistically associated with a reduction in deaths (15). In this initiative, components of the SST (e.g., tension wire sensors) were integrated into a physical measure installed at one bridge that partially restricted access to means. Unlike the physical mitigations deployed in the aforementioned studies (21,29) the installation of these sensors (which were also topped with a spinning rail) would not prevent an individual climbing through them, but would alert officials of any attempts to climb through or cut them. From this perspective, such a mitigation may help to “restrict access to means of suicide” and also “increase opportunity for human intervention”. However, as the study findings were limited to deaths only, it is not possible to confirm the latter. While these data may not always be collected or straightforward to obtain, there may be benefits in reporting on alternative outcomes (e.g., interventions) in studies where surveillance technologies are part of the intervention.

Three studies compared the characteristics of multiple locations across transportation networks, and as such, did not focus exclusively on ‘high-frequency’ locations for suicide or trespass (18–20). In two of these studies, the presence of surveillance technology at subway stations (after controlling for other environmental factors) was not associated with significant differences in rates of suicide attempts or deaths by suicides compared to stations without surveillance technology (18,19). However, another study found that, after controlling for other factors, the number of surveillance “units” identified at railway stations and station car parks within a neighbourhood was negatively associated with rail suicide rates (20). Accounting for the (perceived) coverage of surveillance (rather than simple presence of a camera) may be useful in this context. Indeed, Uittenbogaard & Ceccato (19) found significant associations between suicide risk at station and factors that may impact perceived visibility, such as the presence of over 20 passengers or walls dividing platforms. Moreover, Torok et al. (24) suggested signs of immediate displacement within a cliff site environment may reflect a willingness to seek out areas with reduced visibility. Further research is therefore needed to better understand whether perceived surveillance and its coverage have an overall impact on suicidal behaviours.

While many of the studies featuring surveillance technologies were designed to initiate or aid human-led interventions, several initiated technology-led responses. Studies which included SSTs designed to initiate motion-activated or dynamic lighting offered no conclusive evidence of preventing suicides. No studies of the impact of audible deterrents (either automated or issued remotely from a control room) on suicide outcomes were found in the search. However, three studies examined the effect on trespass (26–28). Although the findings from these studies suggest that SSTs that issue audible deterrents may be helpful for reducing trespass on the railways, their application for suicide prevention is unclear. While further research is required, there is some initial evidence to suggest that perceived risks and consequences may have played some role in determining the effectiveness of audible deterrents at reducing trespass. Indeed, da Silva et al. (26) found that the effectiveness of an SST installed at one rail-bridge location waned over time, potentially due to individuals becoming desensitised to the announcement and awareness that a human-led response was unlikely to arrive. Additionally, Kallberg & Silla (27) found that the system was more effective at reducing trespass when deployed at a site with a higher frequency of trains, that also travelled faster (i.e., a higher-risk site). More broadly, other studies have found that a combination of education on rail safety and threat of punishment may help to prevent unsafe behaviours at level crossings (30,31). In Barić et al., (30) unsafe behaviours were also not influenced by installing CCTV alone. As such the findings may not necessarily translate to suicide prevention. Although some suicide attempts may involve a degree of trespass, the intended outcome is arguably very different from someone who, for example, intends to pass through the environment as a shortcut. Given that train frequency and speed have previously been associated with risk of rail suicide at a location (20,32,33), perceived risk and consequences may be less likely to reliably act as preventive factors in the context of suicide (vs. general trespass).

There were several limitations around the evidence identified in this review. A key limitation is that we only included evidence published in English. As identified in this review, much of the work on SSTs in suicide prevention takes place in countries where English is not the official language. Future research in this area may therefore benefit from international collaboration. Second, while fifteen studies were identified, three examined one suicide prevention project at different points in time (22–24), and potentially three studies looked at different projects installed on the same bridge (15–17). Our knowledge about surveillance technologies is therefore limited to a small number of sites. Moreover, we found SSTs were frequently installed alongside a package of other measures. This means their contribution is often difficult to gauge. In addition, descriptions of SST capabilities and surrounding processes are often limited or absent. Other challenges associated with working with real-world data, such as data availability (e.g., of additional outcomes) and the relative rarity of an event such as suicide (e.g., meaning that certain inferential tests are not always feasible) are another limitation of the evidence in this area.

### Implications for practice, policy and future research

Findings from this review highlight critical gaps in the current evidence. While our initial searches and previous work identified a broader and often more advanced range of surveillance technologies being proposed and deployed for suicide prevention (34) the mechanisms for detection used by surveillance technologies in the studies identified here were relatively limited in comparison (e.g., motion detection), or non-existent (e.g., CCTV). Whether the findings would therefore extend to other detection approaches is unclear. It does, however, raise important questions around the ethics of using “smart” surveillance technologies for suicide prevention in public spaces without evidence not only supporting their effectiveness, but also cost-effectiveness. Further efforts to understand what people with lived experience feel about their use is also vital (35).

The evidence around whether SSTs may have a direct impact on preventing suicides is currently limited. Well-designed natural experimental studies, comparing trends in larger samples of matched intervention and control sites are needed. Moreover, only one study found a significant reduction in suicides following the installation of an SST, and this was notably the one that also included partially restricted access to means via the use of tension-wire sensors (15). Understanding how SSTs may be integrated with physical mitigations to aid suicide prevention efforts may therefore be a useful avenue for future research.

While there was also some evidence to suggest that fully-automated audible deterrents may be effective for reducing trespass rates (27,28), no studies explored whether they would also help to prevent suicides. An important consideration is whether any perceived risks that may deter a trespasser from taking a shortcut (e.g., serious injury) after hearing an automated warning are likely to apply to individuals motivated to make a suicide attempt. Indeed, previous research has highlighted a potential risk that some individuals may move to act more quickly during a suicide attempt if they perceive an imminent intervention (23,36). Others have raised concerns that some SSTs may draw attention to a site (34). As such, the use of fully automated SSTs (e.g., audible deterrents) is not currently recommended for use in suicide prevention based on the lack of evidence found here.

Our findings also suggest that the way in which surveillance technology feeds into wider processes surrounding suicide prevention is an important consideration for practitioners and researchers. In many of the studies identified, surveillance technologies were instead part of the intervention process supporting humans to identify events and initiate a response. From this perspective, SSTs are only valuable for suicide prevention if the processes in place to response are sufficient. Yet, our understanding of how SSTs may indirectly help to prevent suicides in this way remains limited. The installation of SSTs developed to send notifications to staff was associated with an increase in interventions in two studies (16,29). However, most other studies did not examine this outcome. Information about the monitoring of systems and any response mechanisms was also limited across studies where a human-led response would be required. Future research in this area should consider what happens following an alert, as technology is only one part of an intervention in this context (e.g., processes, response times). An SST may be able to detect a person in crisis, for example, but the time it then takes a rescue response to be organised and then arrive at a site may inform whether the technology can actually help to prevent suicides in the real world. Evaluating SSTs against multiple outcomes (e.g., other suicidal behaviours, average response times) may therefore be valuable for understanding both their effectiveness at preventing suicides and the implications for their use for those working on the ground.

## Data Availability

All data produced in the present study are available upon reasonable request to the authors

## Acknowledgments

We would firstly like to thank the group of NSPA (National Suicide Prevention Alliance) Lived Experience Influencers who formed an advisory group that continues to provide invaluable feedback across this project. We are also grateful for the advice and support from our colleagues from the wider project team (consisting of Alex Dark, Amina Mahmood, Anthony Purvis, Carlisle George, Elizabeth Pettersen, Giorgio Ciminata, Ian Marsh, Manuela Deidda, Philip Worrall and Robin Pharoah) and steering group (consisting of Rachel Aldred, Amy Chandler, Frank De Vocht, Keith Hawton, Fiona Malpass, Emma McIntosh, and Hilary Norman). For the purpose of open access, the authors have applied a Creative Commons Attribution (CC BY) licence to any Author Accepted Manuscript version arising from this submission.

## Contributors

Conceptualisation, J.-M.M. and L.M.; Methodology, L.J., J.-M.M., K.H., L.M. and P.C.; Investigation, L.J. and B.C.; Formal analysis, L.J. and B.C.; Writing – Original Draft, L.J.; Writing – Review & Editing, B.C.., J.-M.M., K.H., L.M. and P.C.; Funding Acquisition, J.-M.M. and L.M.

