## Appendix A-C for "The effectiveness of surveillance technology for the prevention of suicides in public spaces: a systematic review"

Search Strategies

**PsycINFO (EBSCO)**

Limiters - English; Language: English, Start year - 1990

| **Terms** |  |
| --- | --- |
| DE( "Suicide" OR "Suicidal Behavior" OR "Suicide Prevention" OR "Suicidology" OR "Attempted Suicide" OR "Nonsuicidal self-injury") | S1 |
| MM( "Suicide" OR "Suicide, Completed" OR "Suicide Prevention" OR "Suicidal Ideation" OR "Suicide, Attempted" OR "Self-Injurious Behavior" ) | S2 |
| TI ( Suicid* OR Self-harm* OR "Near-lethal self-injur*" OR "Near-lethal attempt" OR Trespass*) OR AB ( Suicid* OR Self-harm* OR "Near-lethal self-injur*" OR "Near-lethal attempt" OR Trespass*) | S3 |
| DE ("Artificial Intelligence" OR "Human Technology Interaction" OR "Computer Vision") | S4 |
| TI( "Surveillance Technolog*" OR "Surveillance systems" OR "Remote monitoring" OR "Monitoring systems" OR comput* N3 systems OR "Video surveillance" OR CCTV OR "Closed Circuit Television" OR Camera N3 systems OR ANPR OR LPR OR Infrared N3 Cameras OR "Infrared sensors" OR Thermal N3 Cameras OR "Artificial Intelligence" OR "Intelligent W3 Surveillance" OR "Automat* N3 System" OR "AI N3 System" OR "Computer vision" OR "Analytic* N3 Camera" OR "AI N3 Camera" OR "Smart Camera" OR "Video N3 Analytic*" OR "Motion detect*" OR "Mov* N3 detect*" OR "Mov* N3 track*" OR "Object detect*" OR "Object track*" OR "Object classif*" OR "Behavio#r* detect*" OR "Behavio#r* recogni*" OR "Behavio#r* analysis" OR "Activity detect*" OR "Activity recogni*" OR "Real-time recognition" OR "Video N3 Recognition" OR "Anomaly detect*" OR "Intrusion detect*" OR Automat* N3 detect* OR "Obstacle detect*" OR "Acoustic detect*" OR Sensor N3 System OR Fibre N3 Sensor OR Fiber N3 Sensor OR "Acoustic Sens*" OR "Pressure Sens*" OR "Fac* recognition" OR "Trajectory analysis" OR "Crowd analysis" OR Virtual W3 Fenc* OR Lidar OR Radar OR Beacons OR "Bluetooth Low Energy" OR Drones OR "Unmanned aerial vehicles" OR UAV OR "Internet of Things" OR IoT OR WiFi OR Wi-Fi)  OR AB( "Surveillance Technolog*" OR "Surveillance systems" OR "Remote monitoring" OR "Monitoring systems" OR comput* N3 systems OR "Video surveillance" OR CCTV OR "Closed Circuit Television" OR Camera N3 systems OR ANPR OR LPR OR Infrared N3 Cameras OR "Infrared sensors" OR Thermal N3 Cameras OR "Artificial Intelligence" OR "Intelligent W3 Surveillance" OR "Automat* N3 System" OR "AI N3 System" OR "Computer vision" OR "Analytic* N3 Camera" OR "AI N3 Camera" OR "Smart Camera" OR "Video N3 Analytic*" OR "Motion detect*" OR "Mov* N3 detect*" OR "Mov* N3 track*" OR "Object detect*" OR "Object track*" OR "Object classif*" OR "Behavio#r* detect*" OR "Behavio#r* recogni*" OR "Behavio#r* analysis" OR "Activity detect*" OR "Activity recogni*" OR "Real-time recognition" OR "Video N3 Recognition" OR "Anomaly detect*" OR "Intrusion detect*" OR Automat* N3 detect* OR "Obstacle detect*" OR "Acoustic detect*" OR Sensor N3 System OR Fibre N3 Sensor OR Fiber N3 Sensor OR "Acoustic Sens*" OR "Pressure Sens*" OR "Fac* recognition" OR "Trajectory analysis" OR "Crowd analysis" OR Virtual W3 Fenc* OR Lidar OR Radar OR Beacons OR "Bluetooth Low Energy" OR Drones OR "Unmanned aerial vehicles" OR UAV OR "Internet of Things" OR IoT OR WiFi OR Wi-Fi) | S5 |
| (S1 OR S2 OR S3) AND (S4 OR S5) | S6 |

**MEDLINE (EBSCO)**

Limiters - English; Language: English, Start year - 1990

| **Terms** |  |
| --- | --- |
| MH ("Suicide+" OR "Suicide Prevention+" OR "Suicide Completed+" OR "Suicide Attempted+" OR "Suicide Ideation+" OR "Self-Injurious Behavior") | S1 |
| TI ( Suicid* OR Self-harm* OR "Near-lethal self-injur*" OR "Near-lethal attempt" OR Trespass*) OR AB ( Suicid* OR Self-harm* OR "Near-lethal self-injur*" OR "Near-lethal attempt" OR Trespass*) | S2 |
| TI( "Surveillance Technolog*" OR "Surveillance systems" OR "Remote monitoring" OR "Monitoring systems" OR comput* N3 systems OR "Video surveillance" OR CCTV OR "Closed Circuit Television" OR Camera N3 systems OR ANPR OR LPR OR Infrared N3 Cameras OR "Infrared sensors" OR Thermal N3 Cameras OR "Artificial Intelligence" OR "Intelligent W3 Surveillance" OR "Automat* N3 System" OR "AI N3 System" OR "Computer vision" OR "Analytic* N3 Camera" OR "AI N3 Camera" OR "Smart Camera" OR "Video N3 Analytic*" OR "Motion detect*" OR "Mov* N3 detect*" OR "Mov* N3 track*" OR "Object detect*" OR "Object track*" OR "Object classif*" OR "Behavio#r* detect*" OR "Behavio#r* recogni*" OR "Behavio#r* analysis" OR "Activity detect*" OR "Activity recogni*" OR "Real-time recognition" OR "Video N3 Recognition" OR "Anomaly detect*" OR "Intrusion detect*" OR Automat* N3 detect* OR "Obstacle detect*" OR "Acoustic detect*" OR Sensor N3 System OR Fibre N3 Sensor OR Fiber N3 Sensor OR "Acoustic Sens*" OR "Pressure Sens*" OR "Fac* recognition" OR "Trajectory analysis" OR "Crowd analysis" OR Virtual W3 Fenc* OR Lidar OR Radar OR Beacons OR "Bluetooth Low Energy" OR Drones OR "Unmanned aerial vehicles" OR UAV OR "Internet of Things" OR IoT OR WiFi OR Wi-Fi)  OR AB( "Surveillance Technolog*" OR "Surveillance systems" OR "Remote monitoring" OR "Monitoring systems" OR comput* N3 systems OR "Video surveillance" OR CCTV OR "Closed Circuit Television" OR Camera N3 systems OR ANPR OR LPR OR Infrared N3 Cameras OR "Infrared sensors" OR Thermal N3 Cameras OR "Artificial Intelligence" OR "Intelligent W3 Surveillance" OR "Automat* N3 System" OR "AI N3 System" OR "Computer vision" OR "Analytic* N3 Camera" OR "AI N3 Camera" OR "Smart Camera" OR "Video N3 Analytic*" OR "Motion detect*" OR "Mov* N3 detect*" OR "Mov* N3 track*" OR "Object detect*" OR "Object track*" OR "Object classif*" OR "Behavio#r* detect*" OR "Behavio#r* recogni*" OR "Behavio#r* analysis" OR "Activity detect*" OR "Activity recogni*" OR "Real-time recognition" OR "Video N3 Recognition" OR "Anomaly detect*" OR "Intrusion detect*" OR Automat* N3 detect* OR "Obstacle detect*" OR "Acoustic detect*" OR Sensor N3 System OR Fibre N3 Sensor OR Fiber N3 Sensor OR "Acoustic Sens*" OR "Pressure Sens*" OR "Fac* recognition" OR "Trajectory analysis" OR "Crowd analysis" OR Virtual W3 Fenc* OR Lidar OR Radar OR Beacons OR "Bluetooth Low Energy" OR Drones OR "Unmanned aerial vehicles" OR UAV OR "Internet of Things" OR IoT OR WiFi OR Wi-Fi) | S3 |
| (S1 OR S2) AND (S3) | S4 |

**Computer Source (EBSCO)**

Limiters -Start year - 1990

| **Terms** |  |
| --- | --- |
| DE ("SUICIDE" OR "SUICIDE & psychology" OR "SUICIDE -- Law & legislation" OR "SUICIDE prevention" OR "SELF-injurious behavior") | S1 |
| TI ( Suicid* OR Self-harm* OR "Near-lethal self-injur*" OR "Near-lethal attempt" OR Trespass*) OR AB ( Suicid* OR Self-harm* OR "Near-lethal self-injur*" OR "Near-lethal attempt" OR Trespass*) | S2 |
| DE ("COMPUTER vision" OR "ARTIFICIAL intelligence" OR "ARTIFICIAL intelligence & ethics" OR "ARTIFICIAL intelligence & society" OR "ARTIFICIAL intelligence in industry" OR "ARTIFICIAL intelligence research" OR "CLOSED-circuit television" OR "TECHNOLOGY safety measures" OR "TECHNOLOGY security measures") | S3 |
| TI( "Surveillance Technolog*" OR "Surveillance systems" OR "Remote monitoring" OR "Monitoring systems" OR comput* N3 systems OR "Video surveillance" OR CCTV OR "Closed Circuit Television" OR Camera N3 systems OR ANPR OR LPR OR Infrared N3 Cameras OR "Infrared sensors" OR Thermal N3 Cameras OR "Artificial Intelligence" OR "Intelligent W3 Surveillance" OR "Automat* N3 System" OR "AI N3 System" OR "Computer vision" OR "Analytic* N3 Camera" OR "AI N3 Camera" OR "Smart Camera" OR "Video N3 Analytic*" OR "Motion detect*" OR "Mov* N3 detect*" OR "Mov* N3 track*" OR "Object detect*" OR "Object track*" OR "Object classif*" OR "Behavio#r* detect*" OR "Behavio#r* recogni*" OR "Behavio#r* analysis" OR "Activity detect*" OR "Activity recogni*" OR "Real-time recognition" OR "Video N3 Recognition" OR "Anomaly detect*" OR "Intrusion detect*" OR Automat* N3 detect* OR "Obstacle detect*" OR "Acoustic detect*" OR Sensor N3 System OR Fibre N3 Sensor OR Fiber N3 Sensor OR "Acoustic Sens*" OR "Pressure Sens*" OR "Fac* recognition" OR "Trajectory analysis" OR "Crowd analysis" OR Virtual W3 Fenc* OR Lidar OR Radar OR Beacons OR "Bluetooth Low Energy" OR Drones OR "Unmanned aerial vehicles" OR UAV OR "Internet of Things" OR IoT OR WiFi OR Wi-Fi)  OR AB( "Surveillance Technolog*" OR "Surveillance systems" OR "Remote monitoring" OR "Monitoring systems" OR comput* N3 systems OR "Video surveillance" OR CCTV OR "Closed Circuit Television" OR Camera N3 systems OR ANPR OR LPR OR Infrared N3 Cameras OR "Infrared sensors" OR Thermal N3 Cameras OR "Artificial Intelligence" OR "Intelligent W3 Surveillance" OR "Automat* N3 System" OR "AI N3 System" OR "Computer vision" OR "Analytic* N3 Camera" OR "AI N3 Camera" OR "Smart Camera" OR "Video N3 Analytic*" OR "Motion detect*" OR "Mov* N3 detect*" OR "Mov* N3 track*" OR "Object detect*" OR "Object track*" OR "Object classif*" OR "Behavio#r* detect*" OR "Behavio#r* recogni*" OR "Behavio#r* analysis" OR "Activity detect*" OR "Activity recogni*" OR "Real-time recognition" OR "Video N3 Recognition" OR "Anomaly detect*" OR "Intrusion detect*" OR Automat* N3 detect* OR "Obstacle detect*" OR "Acoustic detect*" OR Sensor N3 System OR Fibre N3 Sensor OR Fiber N3 Sensor OR "Acoustic Sens*" OR "Pressure Sens*" OR "Fac* recognition" OR "Trajectory analysis" OR "Crowd analysis" OR Virtual W3 Fenc* OR Lidar OR Radar OR Beacons OR "Bluetooth Low Energy" OR Drones OR "Unmanned aerial vehicles" OR UAV OR "Internet of Things" OR IoT OR WiFi OR Wi-Fi) | S4 |
| (S1 OR S2) AND (S3 OR S4) | S5 |

**CINAHL (EBSCO)**

Limiters - year - 1990

| **ID** | **Terms** |
| --- | --- |
| S1 | MM("Suicide" OR "Suicide Prevention" OR "Suicide, Attempted" OR "Self-Injurious Behavior") |
| S2 | TI ( Suicid* OR Self-harm* OR "Near-lethal self-injur*" OR "Near-lethal attempt" OR Trespass*) OR AB ( Suicid* OR Self-harm* OR "Near-lethal self-injur*" OR "Near-lethal attempt" OR Trespass*) |
| S3 | MM ("Artificial Intelligence") |
| S4 | TI( "Surveillance Technolog*" OR "Surveillance systems" OR "Remote monitoring" OR "Monitoring systems" OR comput* N3 systems OR "Video surveillance" OR CCTV OR "Closed Circuit Television" OR Camera N3 systems OR ANPR OR LPR OR Infrared N3 Cameras OR "Infrared sensors" OR Thermal N3 Cameras OR "Artificial Intelligence" OR "Intelligent W3 Surveillance" OR "Automat* N3 System" OR "AI N3 System" OR "Computer vision" OR "Analytic* N3 Camera" OR "AI N3 Camera" OR "Smart Camera" OR "Video N3 Analytic*" OR "Motion detect*" OR "Mov* N3 detect*" OR "Mov* N3 track*" OR "Object detect*" OR "Object track*" OR "Object classif*" OR "Behavio#r* detect*" OR "Behavio#r* recogni*" OR "Behavio#r* analysis" OR "Activity detect*" OR "Activity recogni*" OR "Real-time recognition" OR "Video N3 Recognition" OR "Anomaly detect*" OR "Intrusion detect*" OR Automat* N3 detect* OR "Obstacle detect*" OR "Acoustic detect*" OR Sensor N3 System OR Fibre N3 Sensor OR Fiber N3 Sensor OR "Acoustic Sens*" OR "Pressure Sens*" OR "Fac* recognition" OR "Trajectory analysis" OR "Crowd analysis" OR Virtual W3 Fenc* OR Lidar OR Radar OR Beacons OR "Bluetooth Low Energy" OR Drones OR "Unmanned aerial vehicles" OR UAV OåR "Internet of Things" OR IoT OR WiFi OR Wi-Fi) OR AB( "Surveillance Technolog*" OR "Surveillance systems" OR "Remote monitoring" OR "Monitoring systems" OR comput* N3 systems OR "Video surveillance" OR CCTV OR "Closed Circuit Television" OR Camera N3 systems OR ANPR OR LPR OR Infrared N3 Cameras OR "Infrared sensors" OR Thermal N3 Cameras OR "Artificial Intelligence" OR "Intelligent W3 Surveillance" OR "Automat* N3 System" OR "AI N3 System" OR "Computer vision" OR "Analytic* N3 Camera" OR "AI N3 Camera" OR "Smart Camera" OR "Video N3 Analytic*" OR "Motion detect*" OR "Mov* N3 detect*" OR "Mov* N3 track*" OR "Object detect*" OR "Object track*" OR "Object classif*" OR "Behavio#r* detect*" OR "Behavio#r* recogni*" OR "Behavio#r* analysis" OR "Activity detect*" OR "Activity recogni*" OR "Real-time recognition" OR "Video N3 Recognition" OR "Anomaly detect*" OR "Intrusion detect*" OR Automat* N3 detect* OR "Obstacle detect*" OR "Acoustic detect*" OR Sensor N3 System OR Fibre N3 Sensor OR Fiber N3 Sensor OR "Acoustic Sens*" OR "Pressure Sens*" OR "Fac* recognition" OR "Trajectory analysis" OR "Crowd analysis" OR Virtual W3 Fenc* OR Lidar OR Radar OR Beacons OR "Bluetooth Low Energy" OR Drones OR "Unmanned aerial vehicles" OR UAV OR "Internet of Things" OR IoT OR WiFi OR Wi-Fi) |
|  | (S1 OR S2) AND (S3 OR S4) |

**OVID** (Journals@Ovid; Ovid Full text journals; Books@OVID; Social Policy & Practice (SPP))

limit 1 to yr="1990 -Current"

**Search**

((Suicid* or Self-harm* or "Near-lethal self-injur*" or "Near-lethal attempt" or Trespass*) and ("Surveillance Technolog*" or "Surveillance systems" or "Remote monitoring" or "Monitoring systems" or (comput* adj3 systems) or "Video surveillance" or CCTV or "Closed Circuit Television" or (Camera adj3 systems) or ANPR or LPR or (Infrared adj3 Cameras) or "Infrared sensors" or (Thermal adj3 Cameras) or "Artificial Intelligence" or (Intelligent adj3 Surveillance) or (Automat* adj3 System) or (AI adj3 System) or "Computer vision" or (Analytic* adj3 Camera) or (AI adj3 Camera) or "Smart Camera" or (Video adj3 Analytic*) or "Motion detect*" or (Mov* adj3 detect*) or (Mov* adj3 track*) or "Object detect*" or "Object track*" or "Object classif*" or "Behavio#r* detect*" or "Behavio#r* recogni*" or "Behavio#r* analysis" or "Activity detect*" or "Activity recogni*" or "Real-time recognition" or (Video adj3 Recognition) or "Anomaly detect*" or "Intrusion detect*" or (Automat* adj3 detect*) or "Obstacle detect*" or "Acoustic detect*" or (Sensor adj3 System) or (Fibre adj3 Sensor) or (Fiber adj3 Sensor) or "Acoustic Sens*" or "Pressure Sens*" or "Fac* recognition" or "Trajectory analysis" or "Crowd analysis" or (Virtual adj3 Fenc*) or Lidar or Radar or Beacons or "Bluetooth Low Energy" or Drones or "Unmanned aerial vehicles" or UAV or "Internet of Things" or IoT or WiFi or Wi-Fi)).ab,ti,kw.

**WEB OF SCIENCE**

Language: English, Start date – 1990-01-01

**Search**

((TI=(Suicid* OR Self-harm* OR "Near-lethal self-injur*" OR "Near-lethal attempt" OR Trespass* ))

OR (AB=(Suicid* OR Self-harm* OR "Near-lethal self-injur*" OR "Near-lethal attempt" OR Trespass* ))) AND ((TI=("Surveillance Technolog*" OR "Surveillance systems" OR "Remote monitoring" OR "Monitoring systems" OR comput* NEAR/3 systems OR "Video surveillance" OR CCTV OR "Closed Circuit Television" OR Camera NEAR/3 systems OR ANPR OR LPR OR Infrared NEAR/3 Cameras OR "Infrared sensors" OR Thermal NEAR/3 Cameras OR "Artificial Intelligence" OR "Intelligent W3 Surveillance" OR "Automat* NEAR/3 System" OR "AI NEAR/3 System" OR "Computer vision" OR "Analytic* NEAR/3 Camera" OR "AI NEAR/3 Camera" OR "Smart Camera" OR "Video NEAR/3 Analytic*" OR "Motion detect*" OR "Mov* NEAR/3 detect*" OR "Mov* NEAR/3 track*" OR "Object detect*" OR "Object track*" OR "Object classif*" OR "Behavior* detect*" OR "Behavior* recogni*" OR "Behavior* analysis" OR "Behaviour* detect*" OR "Behaviour* recogni*" OR "Behaviour* analysis" OR "Activity detect*" OR "Activity recogni*" OR "Real-time recognition" OR "Video NEAR/3 Recognition" OR "Anomaly detect*" OR "Intrusion detect*" OR Automat* NEAR/3 detect* OR "Obstacle detect*" OR "Acoustic detect*" OR Sensor NEAR/3 System OR Fibre NEAR/3 Sensor OR Fiber NEAR/3 Sensor OR "Acoustic Sens*" OR "Pressure Sens*" OR "Fac* recognition" OR "Trajectory analysis" OR "Crowd analysis" OR Virtual NEAR/3 Fenc* OR Lidar OR Radar OR Beacons OR "Bluetooth Low Energy" OR Drones OR "Unmanned aerial vehicles" OR UAV OR "Internet of Things" OR IoT OR WiFi OR Wi-Fi))

OR (AB=("Surveillance Technolog*" OR "Surveillance systems" OR "Remote monitoring" OR "Monitoring systems" OR comput* NEAR/3 systems OR "Video surveillance" OR CCTV OR "Closed Circuit Television" OR Camera NEAR/3 systems OR ANPR OR LPR OR Infrared NEAR/3 Cameras OR "Infrared sensors" OR Thermal NEAR/3 Cameras OR "Artificial Intelligence" OR "Intelligent W3 Surveillance" OR "Automat* NEAR/3 System" OR "AI NEAR/3 System" OR "Computer vision" OR "Analytic* NEAR/3 Camera" OR "AI NEAR/3 Camera" OR "Smart Camera" OR "Video NEAR/3 Analytic*" OR "Motion detect*" OR "Mov* NEAR/3 detect*" OR "Mov* NEAR/3 track*" OR "Object detect*" OR "Object track*" OR "Object classif*" OR "Behavior* detect*" OR "Behavior* recogni*" OR "Behavior* analysis" OR "Behaviour* detect*" OR "Behaviour* recogni*" OR "Behaviour* analysis" OR "Activity detect*" OR "Activity recogni*" OR "Real-time recognition" OR "Video NEAR/3 Recognition" OR "Anomaly detect*" OR "Intrusion detect*" OR Automat* NEAR/3 detect* OR "Obstacle detect*" OR "Acoustic detect*" OR Sensor NEAR/3 System OR Fibre NEAR/3 Sensor OR Fiber NEAR/3 Sensor OR "Acoustic Sens*" OR "Pressure Sens*" OR "Fac* recognition" OR "Trajectory analysis" OR "Crowd analysis" OR Virtual NEAR/3 Fenc* OR Lidar OR Radar OR Beacons OR "Bluetooth Low Energy" OR Drones OR "Unmanned aerial vehicles" OR UAV OR "Internet of Things" OR IoT OR WiFi OR Wi-Fi)))

**PTSD Pubs**

After 01 January 1990, Search: Anywhere

**Search**

(suicid* OR "Attempted suicide" OR "Completed suicide" OR "Suicide prevention" OR "Suicidal Ideation" OR Self-harm* OR "Near-lethal self-injur*" OR "Near-lethal attempt" OR Trespass*) AND ("artificial intelligence" OR "Human technology interaction" OR "computer vision" OR "surveillance tech*" OR "Surveillance system" OR "remote monitoring" OR "monitoring system" OR "comput* NEAR system*" OR "video surveillance" OR CCTV OR "Closed circuit television" OR "camera NEAR system*" OR ANPR OR LPR OR "Infrared NEAR camera*" OR "infrared sensor*" OR "Thermal NEAR camera*" OR "intelligent surveillance" OR "automat* NEAR system*" OR "AI NEAR system*" OR "computer vision" OR "analytic camera" OR "AI NEAR camera" "Smart Camera" OR "video NEAR analytic*" OR "motion detect*" OR "movement NEAR detect*" OR "Movement NEAR track*" OR "object track*" OR "object detect*" OR "object classif*" OR "behavior detection" OR "behavior recognition" OR "behavior analysis" OR "activity detect*" OR "activity recogni*" OR "real-time recogni*" OR "video recogni*" OR "anomaly detect*" OR "intrusion detect*" OR "automat* NEAR detect*" OR "obstacle detect*" OR "acoustic detect*" OR "Sensor system" OR "fibre NEAR sensor" OR "face recogni*" OR "facial recogni*" OR "trajectory analysis" OR "crowd analys*" OR "virtual NEAR fenc*" OR Lidar OR radar OR beacons OR "bluetooth low energy" OR drones OR "unmanned aerial vehicles" OR UAV OR "Internet of things" OR Wifi OR Wi-fi)

**CENTRAL (Cochrane)**

|  | **Search** |
| --- | --- |
| 1  2  3  4 | MeSH descriptor: [Suicide] explode all trees  MeSH descriptor: [Suicide Prevention] explode all trees  MeSH descriptor: [Suicide, Attempted] explode all trees  MeSH descriptor: [Self-Injurious Behavior] explode all trees |
| 5 | (Self-harm*):ti,ab,kw OR (Near-lethal NEXT self-injur*):ti,ab,kw OR (Near-lethal NEXT attempt):ti,ab,kw OR (Trespass*):ti,ab,kw OR (Suicid*):ti,ab,kw |
| 6 | (Surveillance NEXT Technolog*):ti,ab,kw OR (Surveillance NEXT systems):ti,ab,kw OR (Remote NEXT monitoring):ti,ab,kw OR (Monitoring NEXT systems):ti,ab,kw OR (comput* NEXT systems):ti,ab,kw |
| 7 | ("Video surveillance"):ti,ab,kw OR ("CCTV"):ti,ab,kw OR ("Closed Circuit Television"):ti,ab,kw OR (Camera NEXT systems):ti,ab,kw OR (ANPR):ti,ab,kw |
| 8 | (LPR):ti,ab,kw OR (Infrared NEAR3 Cameras):ti,ab,kw OR ("Infrared sensors"):ti,ab,kw OR (Thermal NEAR3 Cameras):ti,ab,kw OR ("Artificial Intelligence"):ti,ab,kw |
| 9 | (Intelligent Surveillance):ti,ab,kw OR (Automat* NEXT System):ti,ab,kw OR (AI NEXT System):ti,ab,kw OR ("Computer vision"):ti,ab,kw OR (Analytic* Camera):ti,ab,kw |
| 10 | (AI NEXT Camera):ti,ab,kw OR (Smart NEXT Camera):ti,ab,kw OR (Video NEXT Analytic*):ti,ab,kw OR (Motion NEXT detect*):ti,ab,kw OR (Mov* NEXT track*):ti,ab,kw |
| 11 | (Object NEXT detect*):ti,ab,kw OR (Object NEXT track*):ti,ab,kw OR (Object NEXT classif*):ti,ab,kw OR (Behavior* NEXT detect*):ti,ab,kw OR (Behaviour* NEXT detect*):ti,ab,kw |
| 12 | (Behavior* NEXT recogni*):ti,ab,kw OR (Behaviour* NEXT recogni*):ti,ab,kw OR (Behavior* NEXT analysis):ti,ab,kw OR (Behaviour* NEXT analysis):ti,ab,kw OR (Activity NEXT detect*):ti,ab,kw |
| 13 | (Activity NEXT recogni*):ti,ab,kw OR ("Real-time recognition"):ti,ab,kw OR (Video NEXT Recognition):ti,ab,kw OR (Anomaly NEXT detect*):ti,ab,kw OR (Intrusion NEXT detect*):ti,ab,kw |
| 14 | (Automat* detect*):ti,ab,kw OR (Obstacle NEXT detect*):ti,ab,kw OR (Acoustic NEXT detect*):ti,ab,kw OR (Fibre NEAR3 Sensor):ti,ab,kw |
| 15 | (Fiber NEAR3 Sensor):ti,ab,kw OR (Acoustic NEXT Sens*):ti,ab,kw OR (Pressure NEXT Sens*):ti,ab,kw OR (Facial NEXT recognition):ti,ab,kw OR ("Trajectory analysis"):ti,ab,kw |
| 16 | ("Crowd analysis"):ti,ab,kw OR (Virtual NEXT Fenc*):ti,ab,kw OR (Lidar):ti,ab,kw OR (Radar):ti,ab,kw OR (Beacons):ti,ab,kw |
| 17 | (Bluetooth):ti,ab,kw OR (Drones):ti,ab,kw OR ("Unmanned aerial vehicles"):ti,ab,kw OR (UAV):ti,ab,kw |
| 18 | (IoT):ti,ab,kw OR (WiFi):ti,ab,kw OR (Wi-Fi):ti,ab,kw |
| 19 | #1 OR #2 OR #3 OR #4 OR #5 |
| 20 | #6 OR #7 OR #8 OR #9 OR #10 OR #11 OR #12 OR #13 OR #14 OR #15 OR #16 OR #17 OR #18 |
| 21 | #19 AND #20 |

**IEEE Explore**

From 1990

**Search**

### ((("Mesh_Terms": "Suicide+" OR "Mesh_Terms": "Suicide Prevention+" OR "Mesh_Terms": "Suicide Completed+" OR "Mesh_Terms": "Suicide Attempted+" OR "Mesh_Terms": "Suicide Ideation+" OR "Mesh_Terms": "Self-Injurious Behavior") OR ("Publication Title":Suicide OR "Publication Title":Self-harm OR "Publication Title":"Near-lethal self-injury" OR "Publication Title":"Near-lethal attempt" OR "Publication Title":Trespass) OR ("Abstract":Suicide OR "Abstract":Self-harm OR "Abstract":"Near-lethal self-injury" OR "Abstract":"Near-lethal attempt" OR "Abstract":Trespass)))

**Appendix B**

Extracted data

|  | **Heading** | **Example multiple choice (where applicable)** |
| --- | --- | --- |
| Paper details | Paper title | - |
|  | Year | - |
|  | Authors | - |
|  | Contact details / Corresponding author | - |
|  | DOI / URL | - |
|  | Abstract | - |
|  | Keywords | - |
|  | Country | - |
| Publication details | Sector of publication | Academia; Industry |
|  | Type | Academic journal paper; Conference Proceedings |
|  | Sponsorship / funding source (where applicable) | - |
|  | Source (e.g. journal name / conference) | - |
|  | **Peer reviewed?** | Yes; No |
|  | Other (any additional information) | - |
| Data | Research Type | Quantitative; Mixed-methods |
|  | Research Aim | - |
|  | Research Design / Evidence | Pre-post; Focus group |
|  | Sample type (where relevant) | - |
|  | Method of obtaining data | - |
|  | Total sample size | - |
|  | Inclusion criteria | - |
|  | Exclusion criteria | - |
|  | Other | - |
| Intervention | Technology “goal” type (most advanced) | No smart monitoring (e.g., CCTV); Detect presence |
|  | Input (sensor type) | CCTV camera (Live); Infrared camera |
|  | Analytics parameters (where relevant) | Detecting behaviour – crouching; Detecting behaviour - lingering |
|  | Output (response produced) | Audible deterrent – alarm; Notification – control room |
|  | Other | - |
| Context | Location (general) | Railways / Underground; Cliff site |
|  | Location (monitored) | Station; Level crossing |
|  | Description of location | - |
|  | Other interventions present? | - |
|  | Period data covers | - |
|  | Duration of data coverage | - |
|  | Other | - |
| Outcomes of interest | Primary | - |
|  | Secondary | - |
|  | Other | - |
| Summary | Key findings | - |
|  | Other | - |
|  | Identified strengths | - |
|  | Identified limitations | - |
|  | Ethical considerations (incl. lived experience involvement | - |
|  | Identified implications for practice / policy | - |
|  | Identified implications for research | - |

**Appendix C**

Quality Assessment of studies using the Mixed Methods Appraisal Tool (MMAT)

| **"Authors Year"** | **Category of study design** | **Methodological quality criteria** | **Responses** | | | **Comments** |
| --- | --- | --- | --- | --- | --- | --- |
|  |  |  | **Yes** | **No** | **Can't Tell** |  |
| Kõlves et al., 2023 | 3. Quantitative nonrandomized | S1. Are there clear research questions? | X |  |  |  |
|  |  | S2. Do the collected data allow to address the research questions? | X |  |  |  |
|  |  | 3.1. Are the participants representative of the target population? |  |  | 1 | Would not be appropriate / expected in this area |
|  |  | 3.2. Are measurements appropriate regarding both the outcome and intervention (or exposure)? | 1 |  |  |  |
|  |  | 3.3. Are there complete outcome data? | 1 |  |  | Majority of the data (2001-18) from Queensland Suicide Register, but for 2019-2021 interim QSR were used (preliminary reports) |
|  |  | 3.4. Are the confounders accounted for in the design and analysis? |  | 1 |  | Cameras installed alongside phones, and so effectiveness of individual components (i.e. cameras) cannot be determined. However, three comparison locations were also analysed |
|  |  | 3.5. During the study period, is the intervention administered (or exposure occurred) as intended? |  |  | 1 |  |
| ***Total items scored*** |  |  | ***2*** | ***1*** | ***2*** |  |
| Lockley et al., 2014 | 3. Quantitative nonrandomized | S1. Are there clear research questions? | X |  |  |  |
|  |  | S2. Do the collected data allow to address the research questions? | X |  |  |  |
|  |  | 3.1. Are the participants representative of the target population? |  |  | 1 | Would not be appropriate / expected in this area |
|  |  | 3.2. Are measurements appropriate regarding both the outcome and intervention (or exposure)? | 1 |  |  |  |
|  |  | 3.3. Are there complete outcome data? | 1 |  |  | Data for confirmed suicides (2001-2011) analysed separately from reported jumping incidents (2006-2012) - this is made clear |
|  |  | 3.4. Are the confounders accounted for in the design and analysis? |  | 1 |  | Intervention involves installation of several different measures. |
|  |  | 3.5. During the study period, is the intervention administered (or exposure occurred) as intended? |  | 1 |  | System adjusted over time |
| ***Total items scored*** |  |  | ***2*** | ***2*** | ***1*** |  |
| Ross et al., 2020 | 3. Quantitative nonrandomized | S1. Are there clear research questions? | X |  |  |  |
|  |  | S2. Do the collected data allow to address the research questions? | X |  |  |  |
|  |  | 3.1. Are the participants representative of the target population? |  |  | 1 | Would not be appropriate / expected in this area |
|  |  | 3.2. Are measurements appropriate regarding both the outcome and intervention (or exposure)? | 1 |  |  |  |
|  |  | 3.3. Are there complete outcome data? | 1 |  |  |  |
|  |  | 3.4. Are the confounders accounted for in the design and analysis? |  | 1 |  | Intervention involves installation of several different measures. However, data from wider postcode also analysed |
|  |  | 3.5. During the study period, is the intervention administered (or exposure occurred) as intended? |  |  | 1 | Based on other sources, the system was adjusted over time (including the addition of sensors and alarms during the 'postintervention' period) |
| ***Total items scored*** |  |  | ***2*** | ***1*** | ***2*** |  |
| Torok et al., 2023 | 3. Quantitative nonrandomized | S1. Are there clear research questions? | X |  |  |  |
|  |  | S2. Do the collected data allow to address the research questions? | X |  |  |  |
|  |  | 3.1. Are the participants representative of the target population? |  |  | 1 | Would not be appropriate / expected in this area |
|  |  | 3.2. Are measurements appropriate regarding both the outcome and intervention (or exposure)? | 1 |  |  |  |
|  |  | 3.3. Are there complete outcome data? | 1 |  |  | <5% of records excluded due to lack of precision in the geographic data |
|  |  | 3.4. Are the confounders accounted for in the design and analysis? |  | 1 |  | Intervention involves installation of several different measures. However, data from wider area also analysed |
|  |  | 3.5. During the study period, is the intervention administered (or exposure occurred) as intended? |  | 1 |  | System adjusted over time |
| ***Total items scored*** |  |  | ***2*** | ***2*** | ***1*** |  |
| Niederkrotenthaler et al., 2012 | 3. Quantitative nonrandomized | S1. Are there clear research questions? | X |  |  |  |
|  |  | S2. Do the collected data allow to address the research questions? | X |  |  |  |
|  |  | 3.1. Are the participants representative of the target population? |  |  | 1 | Would not be appropriate / expected in this area |
|  |  | 3.2. Are measurements appropriate regarding both the outcome and intervention (or exposure)? | 1 |  |  |  |
|  |  | 3.3. Are there complete outcome data? | 1 |  |  |  |
|  |  | 3.4. Are the confounders accounted for in the design and analysis? | 1 |  |  |  |
|  |  | 3.5. During the study period, is the intervention administered (or exposure occurred) as intended? |  |  | 1 | Only 'presence' of surveillance unit during time period recorded |
| ***Total items scored*** |  |  | ***3*** | ***0*** | ***2*** |  |
| Too et al., 2015 | 3. Quantitative nonrandomized | S1. Are there clear research questions? | X |  |  |  |
|  |  | S2. Do the collected data allow to address the research questions? | X |  |  |  |
|  |  | 3.1. Are the participants representative of the target population? |  |  | 1 | Would not be appropriate / expected in this area |
|  |  | 3.2. Are measurements appropriate regarding both the outcome and intervention (or exposure)? | 1 |  |  |  |
|  |  | 3.3. Are there complete outcome data? | 1 |  |  |  |
|  |  | 3.4. Are the confounders accounted for in the design and analysis? | 1 |  |  |  |
|  |  | 3.5. During the study period, is the intervention administered (or exposure occurred) as intended? |  |  | 1 | Only 'presence' of surveillance unit during time period recorded |
| ***Total items scored*** |  |  | ***3*** | ***0*** | ***2*** |  |
| Uittenbogaard & Ceccato, 2015 | 3. Quantitative nonrandomized | S1. Are there clear research questions? | X |  |  |  |
|  |  | S2. Do the collected data allow to address the research questions? | X |  |  |  |
|  |  | 3.1. Are the participants representative of the target population? |  |  | 1 | Would not be appropriate / expected in this area |
|  |  | 3.2. Are measurements appropriate regarding both the outcome and intervention (or exposure)? | 1 |  |  |  |
|  |  | 3.3. Are there complete outcome data? | 1 |  |  |  |
|  |  | 3.4. Are the confounders accounted for in the design and analysis? | 1 |  |  | As other factors included in analysis, CCTV ends up being excluded from city centre analysis due to multicollinearity |
|  |  | 3.5. During the study period, is the intervention administered (or exposure occurred) as intended? |  |  | 1 | Only 'presence' of CCTV during time period recorded |
| ***Total items scored*** |  |  | ***3*** | ***0*** | ***2*** |  |
| Kim et al., 2019 | 3. Quantitative nonrandomized | S1. Are there clear research questions? | X |  |  |  |
|  |  | S2. Do the collected data allow to address the research questions? | X |  |  |  |
|  |  | 3.1. Are the participants representative of the target population? |  |  | 1 | Would not be appropriate / expected in this area |
|  |  | 3.2. Are measurements appropriate regarding both the outcome and intervention (or exposure)? |  |  | 1 |  |
|  |  | 3.3. Are there complete outcome data? |  |  | 1 |  |
|  |  | 3.4. Are the confounders accounted for in the design and analysis? |  | 1 |  | Overlap with installation of another intervention at same location (i.e. Lee et al. 2016) |
|  |  | 3.5. During the study period, is the intervention administered (or exposure occurred) as intended? |  |  | 1 |  |
| ***Total items scored*** |  |  | ***0*** | ***1*** | ***4*** |  |
| Lee et al., 2016 | 3. Quantitative nonrandomized | S1. Are there clear research questions? | X |  |  |  |
|  |  | S2. Do the collected data allow to address the research questions? | X |  |  |  |
|  |  | 3.1. Are the participants representative of the target population? |  |  | 1 | Would not be appropriate / expected in this area |
|  |  | 3.2. Are measurements appropriate regarding both the outcome and intervention (or exposure)? |  |  | 1 | Not clearly defined (e.g. what constitutes a 'rescue on bridge') |
|  |  | 3.3. Are there complete outcome data? |  |  | 1 | Based on 'operation results' |
|  |  | 3.4. Are the confounders accounted for in the design and analysis? |  | 1 |  | Overlap with installation of another intervention at same location (i.e. Kim et al. 2019) |
|  |  | 3.5. During the study period, is the intervention administered (or exposure occurred) as intended? |  | 1 |  | System adjusted throughout pilot year (for optimisation) |
| ***Total items scored*** |  |  | ***0*** | ***2*** | ***3*** |  |
| Erlangsen et al., 2023 | 3. Quantitative nonrandomized | S1. Are there clear research questions? | X |  |  |  |
|  |  | S2. Do the collected data allow to address the research questions? | X |  |  |  |
|  |  | 3.1. Are the participants representative of the target population? |  |  | 1 | Would not be appropriate / expected in this area |
|  |  | 3.2. Are measurements appropriate regarding both the outcome and intervention (or exposure)? | 1 |  |  |  |
|  |  | 3.3. Are there complete outcome data? | 1 |  |  | Based on data collected by railways |
|  |  | 3.4. Are the confounders accounted for in the design and analysis? |  | 1 |  | Intervention involves installation of several different measures. |
|  |  | 3.5. During the study period, is the intervention administered (or exposure occurred) as intended? |  |  | 1 |  |
| ***Total items scored*** |  |  | ***2*** | ***1*** | ***2*** |  |
| da Silva et al., 2006 | 3. Quantitative nonrandomized | S1. Are there clear research questions? | X |  |  |  |
|  |  | S2. Do the collected data allow to address the research questions? | X |  |  |  |
|  |  | 3.1. Are the participants representative of the target population? |  |  | 1 | Would not be appropriate / expected in this area |
|  |  | 3.2. Are measurements appropriate regarding both the outcome and intervention (or exposure)? | 1 |  |  |  |
|  |  | 3.3. Are there complete outcome data? | 1 |  |  | Locally recorded video reviewed and records added alongside security logs (e.g.. where system had not activated). Data was collected for 129 common days each year (where system was operational across all three years). |
|  |  | 3.4. Are the confounders accounted for in the design and analysis? |  | 1 |  | Authors note there may be potential media exposure effects as the project was publicised |
|  |  | 3.5. During the study period, is the intervention administered (or exposure occurred) as intended? |  | 1 |  | In some instances the announcement procedure was not followed by the security team (e.g. if they could not see trespassers in footage). System was also operational only 74% of the time. |
| ***Total items scored*** |  |  | ***2*** | ***2*** | ***1*** |  |
| Kallberg & Silla, 2017 | 3. Quantitative nonrandomized | S1. Are there clear research questions? | X |  |  |  |
|  |  | S2. Do the collected data allow to address the research questions? | X |  |  |  |
|  |  | 3.1. Are the participants representative of the target population? |  |  | 1 | Would not be appropriate / expected in this area |
|  |  | 3.2. Are measurements appropriate regarding both the outcome and intervention (or exposure)? | 1 |  |  |  |
|  |  | 3.3. Are there complete outcome data? | 1 |  |  |  |
|  |  | 3.4. Are the confounders accounted for in the design and analysis? |  | 1 |  | Authors highlight this limitation in the methods section |
|  |  | 3.5. During the study period, is the intervention administered (or exposure occurred) as intended? |  | 1 |  | One week break in data collection at one site due to SST fault |
| ***Total items scored*** |  |  | ***2*** | ***2*** | ***1*** |  |
| Van Overmeiren, 2019 | 3. Quantitative nonrandomized | S1. Are there clear research questions? |  |  | X | Findings from slides of industry presentation - the level of detail included about research questions, data, etc not sufficient to make quality assessment |
|  |  | S2. Do the collected data allow to address the research questions? |  |  | X |  |
| ***N/A*** |  |  | ***N/A*** | ***N/A*** | ***N/A*** |  |
| Shin, Pirkis, Clapperton et al., 2024 | 3. Quantitative nonrandomized | S1. Are there clear research questions? | X |  |  |  |
|  |  | S2. Do the collected data allow to address the research questions? | X |  |  |  |
|  |  | 3.1. Are the participants representative of the target population? |  |  | 1 | Would not be appropriate / expected in this area |
|  |  | 3.2. Are measurements appropriate regarding both the outcome and intervention (or exposure)? | 1 |  |  |  |
|  |  | 3.3. Are there complete outcome data? | 1 |  |  |  |
|  |  | 3.4. Are the confounders accounted for in the design and analysis? |  | 1 |  | Authors highlight this as a limitation in the study |
|  |  | 3.5. During the study period, is the intervention administered (or exposure occurred) as intended? |  |  | 1 |  |
| ***Total items scored*** |  |  | ***2*** | ***1*** | ***2*** |  |
| Shin, Pirkis, Spittal et al., 2024 | 3. Quantitative nonrandomized | S1. Are there clear research questions? | X |  |  |  |
|  |  | S2. Do the collected data allow to address the research questions? | X |  |  |  |
|  |  | 3.1. Are the participants representative of the target population? |  |  | 1 | Would not be appropriate / expected in this area |
|  |  | 3.2. Are measurements appropriate regarding both the outcome and intervention (or exposure)? | 1 |  |  |  |
|  |  | 3.3. Are there complete outcome data? | 1 |  |  |  |
|  |  | 3.4. Are the confounders accounted for in the design and analysis? |  | 1 |  | Not possible to know what happened in wider area, however, the introduction of different measures were staggered which is one strength here. |
|  |  | 3.5. During the study period, is the intervention administered (or exposure occurred) as intended? |  |  | 1 |  |
| ***Total items scored*** |  |  | ***2*** | ***1*** | ***2*** |  |
